# Upper cervical spinal cord morphometric charts: from population references to individualized clinical assessment

**DOI:** 10.64898/2026.10.09.26364862

**Authors:** Jiajia Wang, Tiantian Hua, Haiyan Wu, Rui Zhang, Siyao Xu, Cong Zhang, Li Chai, Yajun Yao, Tianyu Gao, Lu Bai, Xiaozhou Du, Chao Zhang, Bingjie Lin, Lifang Liao, Huiying Wang, Yuqi Luo, Xiaoqin Zhu, Demao Deng, Yuankui Wu, Renlong Zhang, Zhiqiang Zhang, Fuqing Zhou, Jiajia Zhu, Xi-Nian Zuo, Shufang Cheng, Xiaona Xia, Xiangshui Meng, Ya-Zhuo Kong, Julien Cohen-Adad, Ruomi Guo, Shuiwei Xia, Yunyun Duan, Zhizheng Zhuo, Yaou Liu, Chinese Brain Health Imaging Consortium (CHART)

## Abstract

The upper cervical spinal cord is essential for motor, sensory and autonomic function, yet its normative morphometric trajectories and disease-related deviations remain unclear. Using three-dimensional T1-weighted images from 10,159 healthy individuals aged 8-85 years, we established normative references for averaged and level-specific upper cervical spinal cord morphometry, revealing measure-specific trajectories with peak ages of 29.7-44.1 years. These references were applied to 2,394 patients with seven neurological disorders: mild cognitive impairment, Alzheimer’s disease, Parkinson’s disease, cerebral small vessel disease, multiple sclerosis, neuromyelitis optica spectrum disorder and myelin oligodendrocyte glycoprotein antibody-associated disease to derive individualized deviation scores. These deviation scores captured disease-specific abnormalities, enabled disease classification, and were associated with cognition, physical disability and disease progression. Finally, a personalized upper cervical spinal cord report was developed to support individualized imaging interpretation. Together, these findings provide a normative framework for quantifying upper cervical spinal cord morphometry and advancing personalized assessment across neurological disorders.

## Introduction

The cervical spinal cord is a critical component of the central nervous system (CNS), mediating motor, sensory and autonomic functions. Upper cervical spinal cord atrophy is a well-documented feature of inflammatory demyelinating diseases, particularly multiple sclerosis (MS)^[1, 2]^ and neuromyelitis optica spectrum disorder (NMOSD)^[2, 3]^. Emerging evidence has also linked such atrophy to cerebral small vessel disease (CSVD)^[4]^ and Alzheimer’s disease (AD)^[5]^, indicating that spinal cord involvement extends to neurodegenerative disorders and cerebrovascular disease. The mean upper cervical cord area (MUCCA), defined as the averaged cross-sectional area (CSA) across the C1-C3 levels, is a widely used surrogate biomarker of upper cervical spinal cord atrophy^[6]^ and shows strong clinical utility, being closely associated with physical disability in MS and NMOSD^[3, 7]^, disease severity in CSVD^[4]^, and cognitive decline in AD^[8]^. Despite this established clinical relevance, MUCCA represents an averaged measure of upper cervical spinal cord morphology and may not fully capture complex spatial and multidimensional patterns of spinal cord alterations. Furthermore, conventional case-control designs characterize average differences between groups but provide limited insight into individual-level variability of upper cervical spinal cord morphology^[9]^.

To address the limitations of conventional group-level comparisons, normative modeling provides a framework for quantifying the extent to which an individual’s morphometric measures deviate from population-based normative references^[10]^. Built on large-scale datasets with advanced harmonization techniques across scanners and sites, this approach enables the estimation of individualized deviation scores^[11, 12]^. Most normative modeling studies have focused on brain structures, in which deviation scores help characterize structural abnormalities and inform differential diagnosis, prognosis, and treatment evaluation^[12–15]^. Despite recent initiatives to build normative MRI biomarker values for the spinal cord^[16, 17]^, spinal cord normative modeling has yet to be widely applied across neurological disorders. In NMOSD, a recent study based on three-dimensional (3D) T1-weighted (T1w) MRI reported that annual spinal cord atrophy rates derived from deviation scores of upper cervical spinal cord morphometry (CSA, antero-posterior diameter [APD], and right-left diameter [RLD]) were associated with disease activity, disease duration, and changes after rituximab treatment^[18]^. This study used measures averaged across the upper cervical spinal cord, leaving the clinical significance of level-specific measures (C1, C2, and C3) across neurological disorders largely unexplored. Furthermore, previous normative modeling studies have primarily focused on brain and upper cervical spinal cord deviations separately, leaving their relationships insufficiently characterized.

In this study, we leveraged a large multicenter dataset of 3D T1w brain MRI scans from 10,159 healthy subjects to establish age- and sex-specific normative references for upper cervical spinal cord morphometry. Morphometric measures, including CSA, APD, and RLD, were quantified both as averaged C1-C3 measures (MUCCA, mean C1-C3 APD and RLD) and as level-specific measures at C1, C2, and C3. Using these references, we characterized individualized deviation patterns in 2,394 patients across seven neurological disorders, including neurodegenerative disorders (mild cognitive impairment [MCI], AD, Parkinson’s disease [PD]), CSVD, and inflammatory demyelinating diseases (MS, NMOSD, and myelin oligodendrocyte glycoprotein antibody-associated disease [MOGAD]). We further examined associations between upper cervical spinal cord deviations and brain structural deviations, and evaluated the clinical utility of upper cervical spinal cord deviations for disease classification, clinical correlation, and prediction of disease progression. Together, these analyses aimed to establish normative references for upper cervical spinal cord morphometry, characterize disease-specific deviation patterns, and explore their potential clinical utility across neurological disorders.

## Results

### Demographic and clinical characteristics

A total of 10,159 HCs aged 8-85 years and 2,394 patients with seven neurological disorders were included (**Fig. S1**). The patient group included patients with neurodegenerative disorders (MCI [n = 157], AD [n = 309], PD [n = 911]), cerebrovascular disease (CSVD [n = 416]) and inflammatory demyelinating diseases (MS [n = 382], AQP4+ NMOSD [n = 171], MOGAD [n = 48]). Demographic and clinical characteristics are summarized in **Table 1**.

**Table 1.**
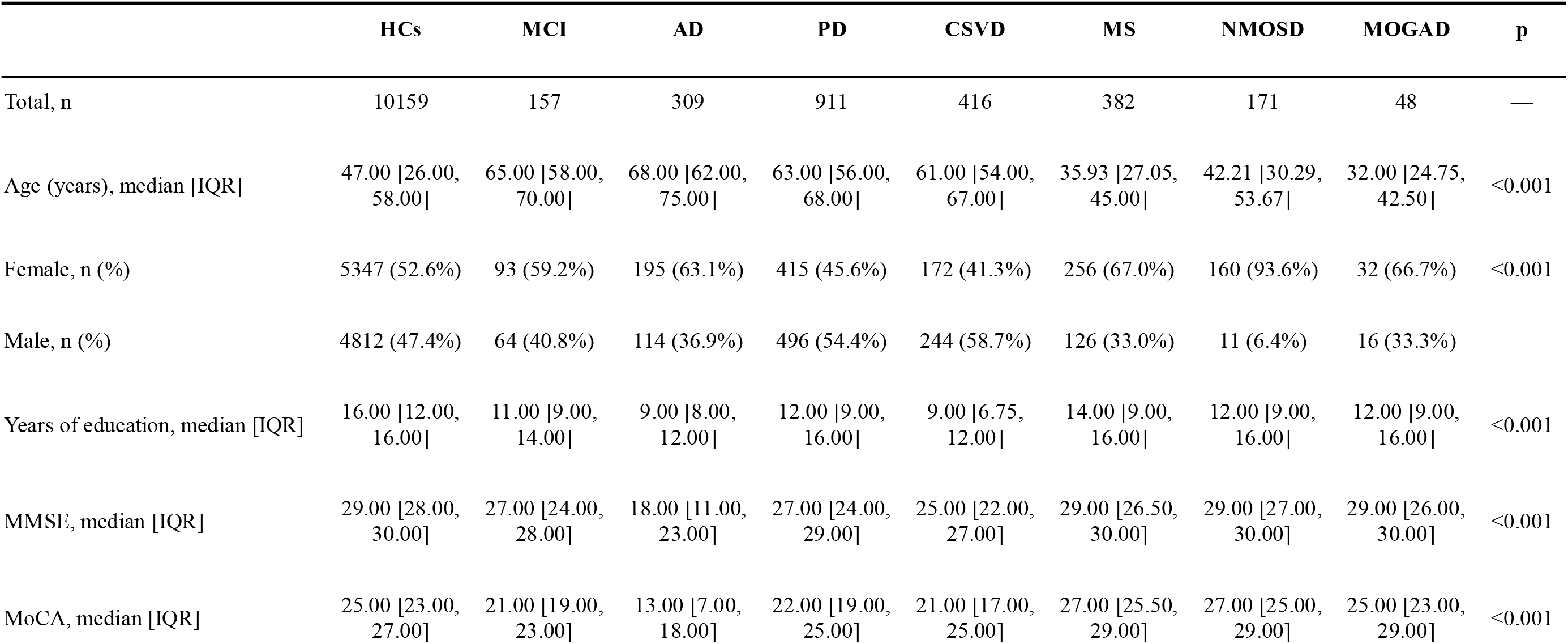
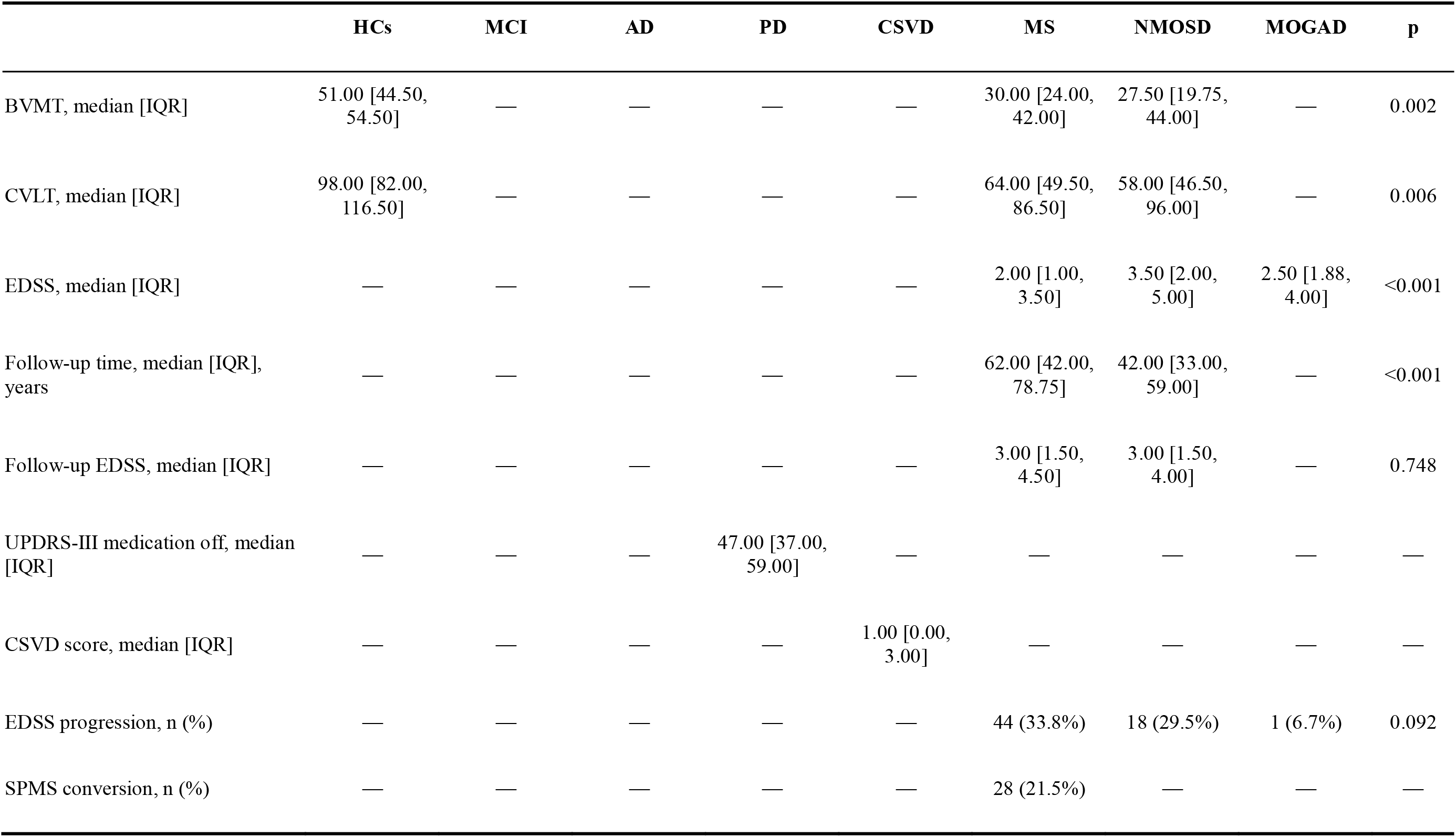
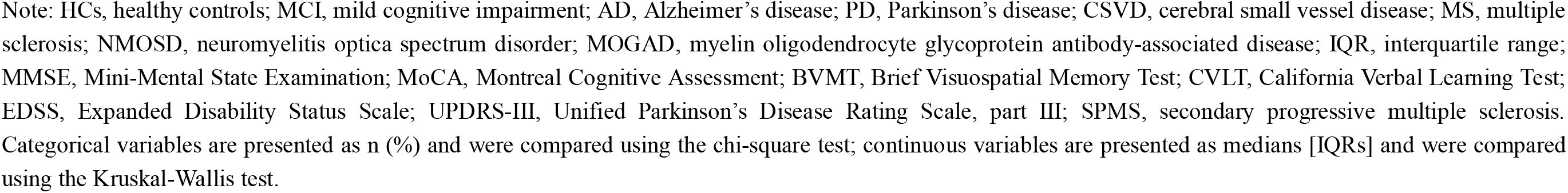
Demographic and clinical characteristics of participants.

|  | HCs | MCI | AD | PD | CSVD | MS | NMOSD | MOGAD | p |
| --- | --- | --- | --- | --- | --- | --- | --- | --- | --- |
| Total, n | 10159 | 157 | 309 | 911 | 416 | 382 | 171 | 48 | — |
| Age (years), median [IQR] | 47.00 [26.00, 58.00] | 65.00 [58.00, 70.00] | 68.00 [62.00, 75.00] | 63.00 [56.00, 68.00] | 61.00 [54.00, 67.00] | 35.93 [27.05, 45.00] | 42.21 [30.29, 53.67] | 32.00 [24.75, 42.50] | <0.001 |
| Female, n (%) | 5347 (52.6%) | 93 (59.2%) | 195 (63.1%) | 415 (45.6%) | 172 (41.3%) | 256 (67.0%) | 160 (93.6%) | 32 (66.7%) | <0.001 |
| Male, n (%) | 4812 (47.4%) | 64 (40.8%) | 114 (36.9%) | 496 (54.4%) | 244 (58.7%) | 126 (33.0%) | 11 (6.4%) | 16 (33.3%) |  |
| Years of education, median [IQR] | 16.00 [12.00, 16.00] | 11.00 [9.00, 14.00] | 9.00 [8.00, 12.00] | 12.00 [9.00, 16.00] | 9.00 [6.75, 12.00] | 14.00 [9.00, 16.00] | 12.00 [9.00, 16.00] | 12.00 [9.00, 16.00] | <0.001 |
| MMSE, median [IQR] | 29.00 [28.00, 30.00] | 27.00 [24.00, 28.00] | 18.00 [11.00, 23.00] | 27.00 [24.00, 29.00] | 25.00 [22.00, 27.00] | 29.00 [26.50, 30.00] | 29.00 [27.00, 30.00] | 29.00 [26.00, 30.00] | <0.001 |
| MoCA, median [IQR] | 25.00 [23.00, 27.00] | 21.00 [19.00, 23.00] | 13.00 [7.00, 18.00] | 22.00 [19.00, 25.00] | 21.00 [17.00, 25.00] | 27.00 [25.50, 29.00] | 27.00 [25.00, 29.00] | 25.00 [23.00, 29.00] | <0.001 |
| BVMT, median [IQR] | 51.00 [44.50, 54.50] | — | — | — | — | 30.00 [24.00, 42.00] | 27.50 [19.75, 44.00] | — | 0.002 |
| CVLT, median [IQR] | 98.00 [82.00, 116.50] | — | — | — | — | 64.00 [49.50, 86.50] | 58.00 [46.50, 96.00] | — | 0.006 |
| EDSS, median [IQR] | — | — | — | — | — | 2.00 [1.00, 3.50] | 3.50 [2.00, 5.00] | 2.50 [1.88, 4.00] | <0.001 |
| Follow-up time, median [IQR], years | — | — | — | — | — | 62.00 [42.00, 78.75] | 42.00 [33.00, 59.00] | — | <0.001 |
| Follow-up EDSS, median [IQR] | — | — | — | — | — | 3.00 [1.50, 4.50] | 3.00 [1.50, 4.00] | — | 0.748 |
| UPDRS-III medication off, median [IQR] | — | — | — | 47.00 [37.00, 59.00] | — | — | — | — | — |
| CSVD score, median [IQR] | — | — | — | — | 1.00 [0.00, 3.00] | — | — | — | — |
| EDSS progression, n (%) | — | — | — | — | — | 44 (33.8%) | 18 (29.5%) | 1 (6.7%) | 0.092 |
| SPMS conversion, n (%) | — | — | — | — | — | 28 (21.5%) | — | — | — |

Note: HCs, healthy controls; MCI, mild cognitive impairment; AD, Alzheimer's disease; PD, Parkinson's disease; CSVD, cerebral small vessel disease; MS, multiple sclerosis; NMOSD, neuromyelitis optica spectrum disorder; MOGAD, myelin oligodendrocyte glycoprotein antibody-associated disease; IQR, interquartile range; MMSE, Mini-Mental State Examination; MoCA, Montreal Cognitive Assessment; BVMC, Brief Visuospatial Memory Test; CVLT, California Verbal Learning Test; EDSS, Expanded Disability Status Scale; UPDRS-III, Unified Parkinson's Disease Rating Scale, part III; SPMS, secondary progressive multiple sclerosis. Categorical variables are presented as n (%) and were compared using the chi-square test; continuous variables are presented as medians [IQRs] and were compared using the Kruskal-Wallis test.

**Table 2.**
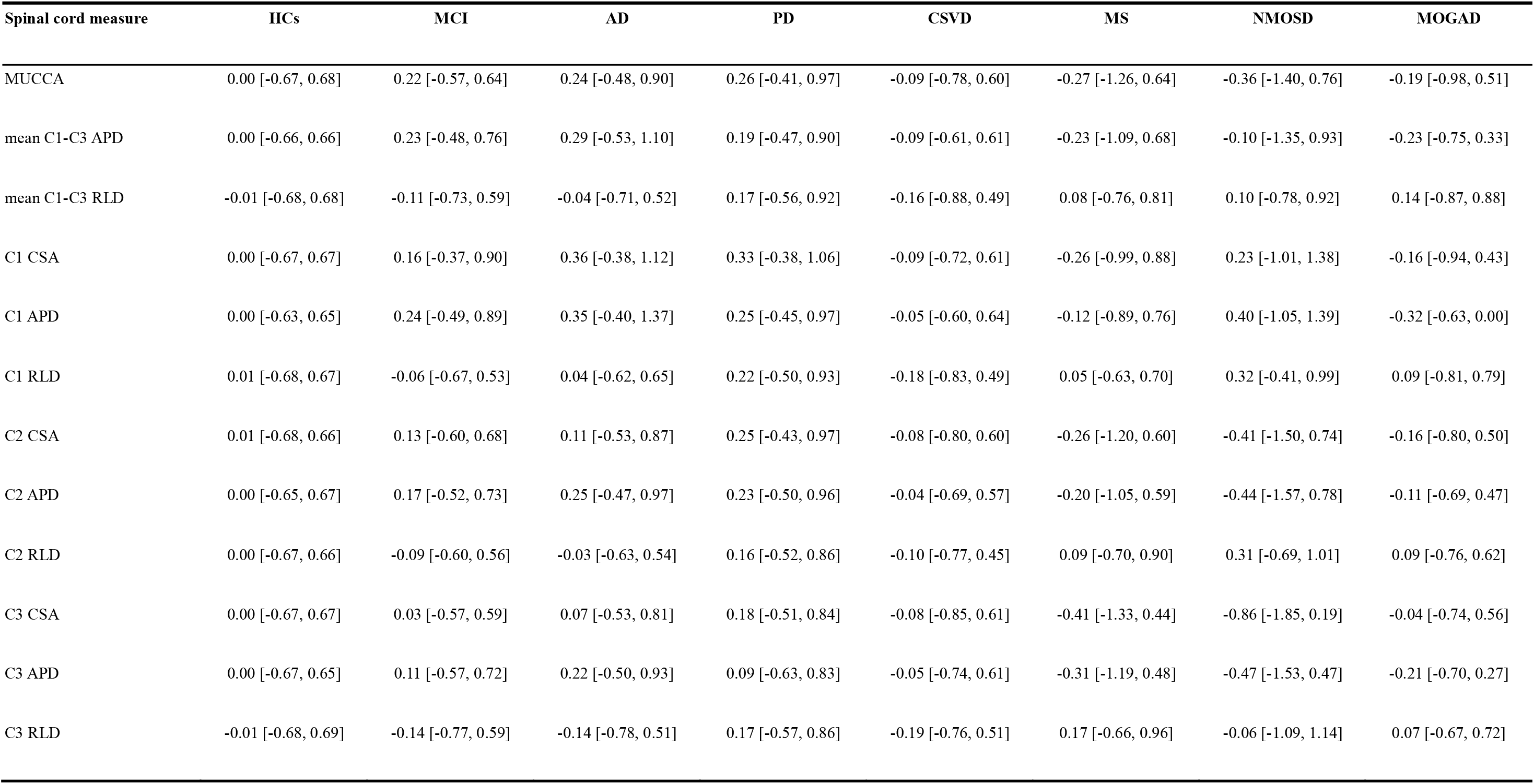
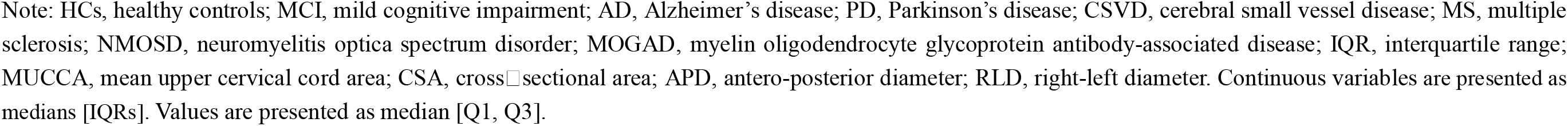
Individual Z scores of upper cervical spinal cord morphometry in HCs and disease groups based on normative reference.

| Spinal cord measure | HCs | MCI | AD | PD | CSVD | MS | NMOSD | MOGAD |
| --- | --- | --- | --- | --- | --- | --- | --- | --- |
| MUCCA | 0.00 [-0.67, 0.68] | 0.22 [-0.57, 0.64] | 0.24 [-0.48, 0.90] | 0.26 [-0.41, 0.97] | -0.09 [-0.78, 0.60] | -0.27 [-1.26, 0.64] | -0.36 [-1.40, 0.76] | -0.19 [-0.98, 0.51] |
| mean C1-C3 APD | 0.00 [-0.66, 0.66] | 0.23 [-0.48, 0.76] | 0.29 [-0.53, 1.10] | 0.19 [-0.47, 0.90] | -0.09 [-0.61, 0.61] | -0.23 [-1.09, 0.68] | -0.10 [-1.35, 0.93] | -0.23 [-0.75, 0.33] |
| mean C1-C3 RLD | -0.01 [-0.68, 0.68] | -0.11 [-0.73, 0.59] | -0.04 [-0.71, 0.52] | 0.17 [-0.56, 0.92] | -0.16 [-0.88, 0.49] | 0.08 [-0.76, 0.81] | 0.10 [-0.78, 0.92] | 0.14 [-0.87, 0.88] |
| C1 CSA | 0.00 [-0.67, 0.67] | 0.16 [-0.37, 0.90] | 0.36 [-0.38, 1.12] | 0.33 [-0.38, 1.06] | -0.09 [-0.72, 0.61] | -0.26 [-0.99, 0.88] | 0.23 [-1.01, 1.38] | -0.16 [-0.94, 0.43] |
| C1 APD | 0.00 [-0.63, 0.65] | 0.24 [-0.49, 0.89] | 0.35 [-0.40, 1.37] | 0.25 [-0.45, 0.97] | -0.05 [-0.60, 0.64] | -0.12 [-0.89, 0.76] | 0.40 [-1.05, 1.39] | -0.32 [-0.63, 0.00] |
| C1 RLD | 0.01 [-0.68, 0.67] | -0.06 [-0.67, 0.53] | 0.04 [-0.62, 0.65] | 0.22 [-0.50, 0.93] | -0.18 [-0.83, 0.49] | 0.05 [-0.63, 0.70] | 0.32 [-0.41, 0.99] | 0.09 [-0.81, 0.79] |
| C2 CSA | 0.01 [-0.68, 0.66] | 0.13 [-0.60, 0.68] | 0.11 [-0.53, 0.87] | 0.25 [-0.43, 0.97] | -0.08 [-0.80, 0.60] | -0.26 [-1.20, 0.60] | -0.41 [-1.50, 0.74] | -0.16 [-0.80, 0.50] |
| C2 APD | 0.00 [-0.65, 0.67] | 0.17 [-0.52, 0.73] | 0.25 [-0.47, 0.97] | 0.23 [-0.50, 0.96] | -0.04 [-0.69, 0.57] | -0.20 [-1.05, 0.59] | -0.44 [-1.57, 0.78] | -0.11 [-0.69, 0.47] |
| C2 RLD | 0.00 [-0.67, 0.66] | -0.09 [-0.60, 0.56] | -0.03 [-0.63, 0.54] | 0.16 [-0.52, 0.86] | -0.10 [-0.77, 0.45] | 0.09 [-0.70, 0.90] | 0.31 [-0.69, 1.01] | 0.09 [-0.76, 0.62] |
| C3 CSA | 0.00 [-0.67, 0.67] | 0.03 [-0.57, 0.59] | 0.07 [-0.53, 0.81] | 0.18 [-0.51, 0.84] | -0.08 [-0.85, 0.61] | -0.41 [-1.33, 0.44] | -0.86 [-1.85, 0.19] | -0.04 [-0.74, 0.56] |
| C3 APD | 0.00 [-0.67, 0.65] | 0.11 [-0.57, 0.72] | 0.22 [-0.50, 0.93] | 0.09 [-0.63, 0.83] | -0.05 [-0.74, 0.61] | -0.31 [-1.19, 0.48] | -0.47 [-1.53, 0.47] | -0.21 [-0.70, 0.27] |
| C3 RLD | -0.01 [-0.68, 0.69] | -0.14 [-0.77, 0.59] | -0.14 [-0.78, 0.51] | 0.17 [-0.57, 0.86] | -0.19 [-0.76, 0.51] | 0.17 [-0.66, 0.96] | -0.06 [-1.09, 1.14] | 0.07 [-0.67, 0.72] |

Note: HCs, healthy controls; MCI, mild cognitive impairment; AD, Alzheimer's disease; PD, Parkinson's disease; CSVD, cerebral small vessel disease; MS, multiple sclerosis; NMOSD, neuromyelitis optica spectrum disorder; MOGAD, myelin oligodendrocyte glycoprotein antibody-associated disease; IQR, interquartile range; MUCCA, mean upper cervical cord area; CSA, cross-sectional area; APD, antero-posterior diameter; RLD, right-left diameter. Continuous variables are presented as medians [IQRs]. Values are presented as median [Q1, Q3].

### Normative modeling of upper cervical spinal cord measures

To establish normative references for upper cervical spinal cord morphometric measures, we collected 3D T1w MRI data from 10,159 HCs aged 8-85 years (median [interquartile range, IQR]: 47.00 [26.00, 58.00] years, 5,347 females [52.6%]) (**Table 1**). Upper cervical spinal cord segmentation was performed using Spinal Cord Toolbox (SCT version 6.0)^[19]^. The extracted morphometric measures included CSA, APD, and RLD, quantified both as averaged C1-C3 measures (MUCCA, mean C1-C3 APD, and RLD), and as level-specific measures at C1, C2, and C3. Normative references were fitted in HCs using generalized additive models for location, scale, and shape (GAMLSS), with each morphometric measure treated as the dependent variable, age and sex included as fixed-effect predictors, and site-specific MRI protocol included as a random-effect term^[12, 15]^. Age effects were modeled non-linearly using fractional polynomial terms, and the optimal model was selected by Bayesian information criterion (BIC). All upper cervical spinal cord morphometric measures exhibited inverted U-shaped age-dependent trajectories across ages 8-85 years (**Fig. 2a**), increasing steadily during development, peaking at 29.7-44.1 years, and gradually decreasing thereafter. The averaged C1-C3 measures showed peak ages within a narrow range of 37.0-38.2 years. Level-specific measures showed comparable peak ages at the C1 and C2 levels, whereas those at the C3 level showed greater variability, with C3 APD peaking at 29.7 years and C3 RLD peaking at 44.1 years (**Fig. 2a**). Cross-sectionally inferred relative rates of change (%/year) showed gradual deceleration prior to the peak age (the age at which the relative rate of change equals zero), followed by increasingly pronounced declines, with median values ranging from 0.094%/year to 0.388%/year before the peak and −0.336%/year to −0.119%/year after the peak. Among the three morphometric dimensions, CSA showed the greatest variability in age-related relative rates of change (standard deviation [SD] range for CSA measures: 0.382-0.528%/year), whereas RLD showed comparatively small variability (SD range for RLD measures: 0.161-0.194%/year) (**Fig. 2b**).

**Fig. 1.**
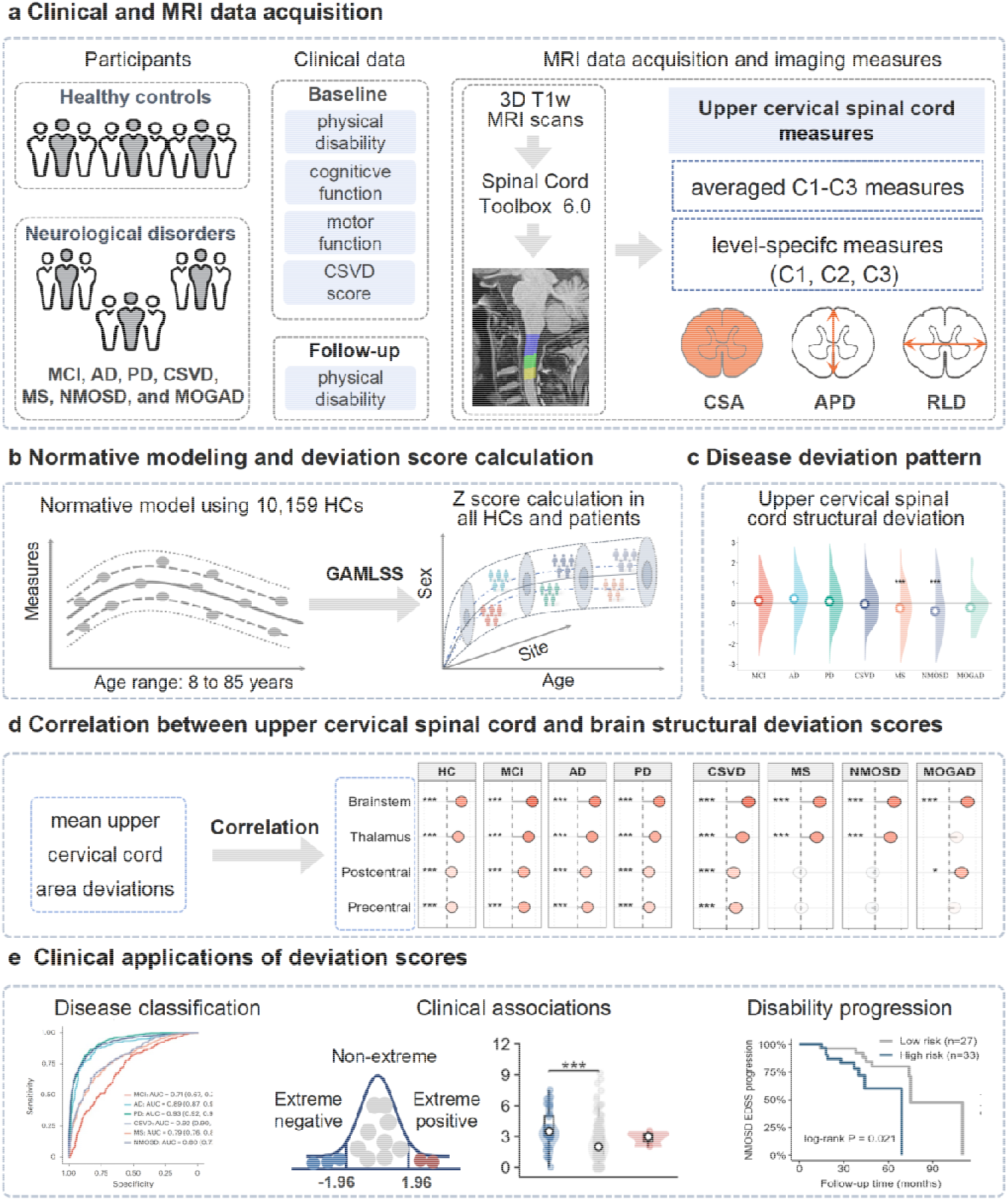
Overview of the study design. (a) Data preparation, including clinical and imaging data from HCs and seven neurological disorders. (b) Imaging segmentation to obtain spinal cord and brain volume measures. (c) Normative modeling of spinal cord structural measures in the healthy population. (d) Calculation of individual deviations in spinal cord structural measures, with adjustment for age, sex and scanning site. (e) Clinical applications of the upper cervical spinal cord deviation scores in neurological disorders. Note: GAMLSS, generalized additive model for location, scale, and shape; CSA, cross sectional area; APD, antero-posterior diameter; RLD, right-left diameter; HCs, healthy controls; MCI, mild cognitive impairment; AD, Alzheimer’s disease; PD, Parkinson’s disease; CSVD, cerebral small vessel disease; MS, multiple sclerosis; NMOSD, neuromyelitis optica spectrum disorder; MOGAD, myelin oligodendrocyte glycoprotein antibody-associated disease.

**Fig. 2.**
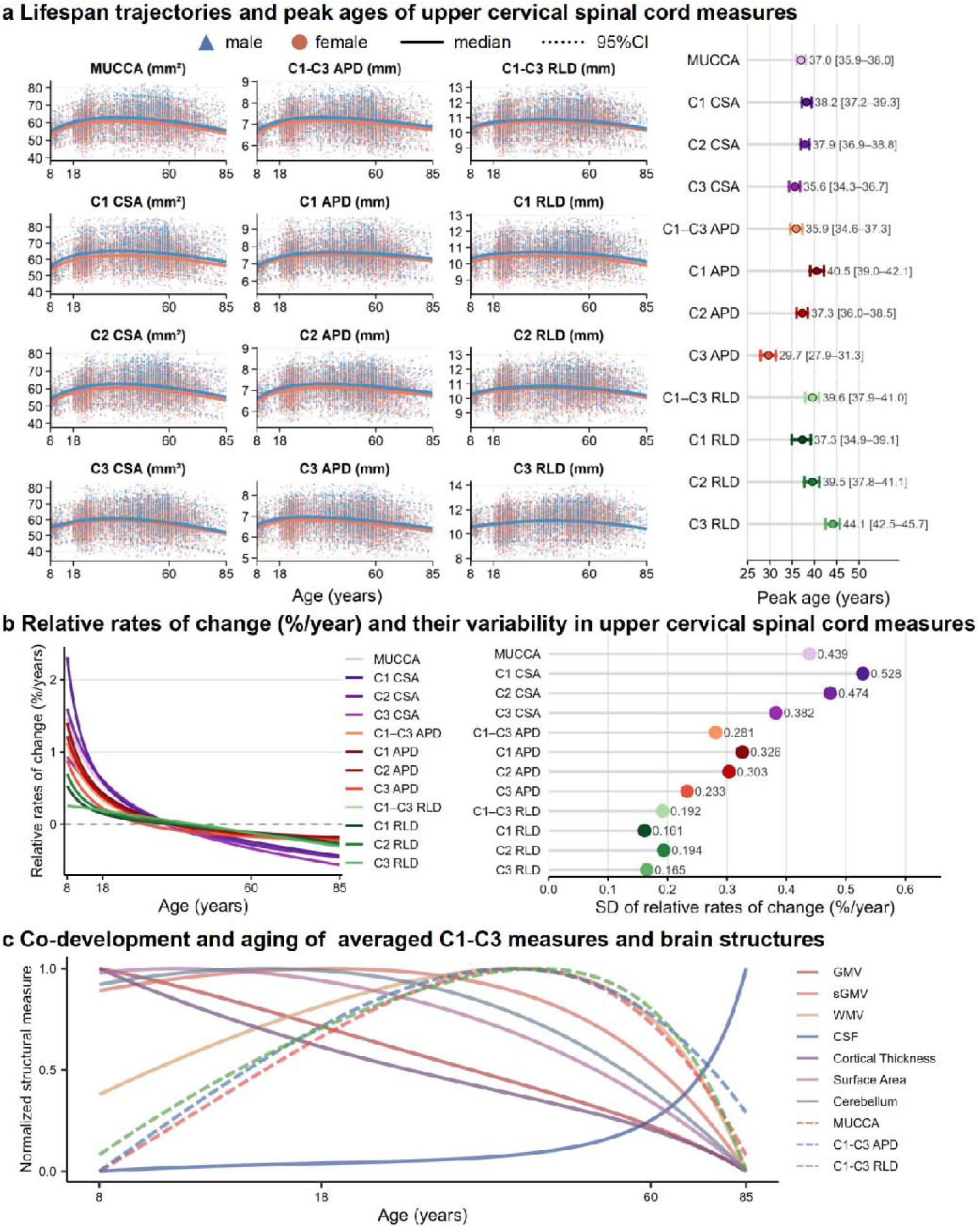
Normative lifespan trajectories of upper cervical spinal cord morphometric measures. (a) Normative trajectories and corresponding peak ages of upper cervical spinal cord morphometric measures across ages 8-85 years. Peak age uncertainty was evaluated by bootstrap resampling (1,000 iterations), with 95% CIs calculated from refitted models. (b) Relative rates of change in upper cervical spinal cord morphometric measures (left panel) and the variability of these rates across the age range, quantified by their SDs (right panel). (c) Normalized trajectories illustrating the co-development and aging patterns of upper cervical spinal cord morphometric measures in relation to brain structural measures. Note: MUCCA, mean upper cervical cord area; CSA, cross sectional area; APD, antero-posterior diameter; RLD, right-left diameter; GMV, gray matter volume; sGMV, subcortical gray matter volume; WMV, white matter volume; CSF, cerebrospinal fluid; SD, standard deviation.

To assess whether age-related changes in the upper cervical spinal cord reflect a developmental and aging pattern shared across the CNS, we compared the normative trajectories of the averaged C1-C3 measures with those of brain morphometric measures, including gray matter volume (GMV), subcortical gray matter volume (sGMV), white matter volume (WMV), cerebrospinal fluid (CSF) volume, cortical thickness, surface area, cerebellum volume. Because the spinal cord participates in ascending and descending sensorimotor pathways, we additionally examined the brainstem, thalamus, postcentral gyrus, and precentral gyrus, which are key components of sensorimotor processing. After rescaling all trajectories to a standardized 0-1 range, the averaged C1-C3 measures showed an inverted U-shaped pattern resembling that of WMV (**Fig. 2c**). Among the sensorimotor-related brain structures, the brainstem displayed a comparable inverted U-shaped trajectory, whereas the thalamus followed an asymmetric inverted U-shaped trajectory, with a modest rise before the peak age and a steeper decline thereafter (**Fig. S2a**). Overall, the age-related trajectory of the upper cervical spinal cord appeared most similar to those of WMV and brainstem volume.

### Upper cervical spinal cord deviations across neurological disorders

Individual upper cervical spinal cord deviation scores (Z scores) were calculated for HCs and patients based on the normative reference framework. For each measure, differences in deviation scores between each patient group and its corresponding age-, sex-, and site-matched subset of HCs were assessed using Monte Carlo permutation tests. Effect sizes were quantified using Cohen’s *d*, with negative values indicating lower deviation scores relative to HCs, and p values were corrected using the false discovery rate (FDR) method.

Across disease groups, upper cervical spinal cord deviation scores showed disease-specific patterns in both the averaged C1-C3 measures and level-specific measures (**Fig. 3a**). At the averaged C1-C3 level, these patterns were characterized by higher values in AD and PD, but lower values in vascular and inflammatory demyelinating diseases. MCI exhibited no significant deviations in any of the three averaged measures (all pFDR > 0.05). AD showed higher values in MUCCA (Cohen’s *d* = 0.20, pFDR = 0.024) and mean C1-C3 APD (Cohen’s *d* = 0.28, pFDR = 0.008), and PD showed higher values across all three averaged measures (Cohen’s *d* = 0.18 to 0.24, all pFDR < 0.01). In contrast, CSVD showed lower values in MUCCA (Cohen’s *d* = −0.14, pFDR = 0.042) and mean C1-C3 RLD (Cohen’s *d* = −0.31, pFDR = 0.031), MS showed lower values in MUCCA (Cohen’s *d* = −0.29, pFDR = 0.003) and mean C1-C3 APD (Cohen’s *d* = −0.18, pFDR = 0.049), NMOSD showed a selective reduction in MUCCA (Cohen’s *d* = −0.29, pFDR < 0.018), whereas MOGAD showed a selective reduction in mean C1-C3 APD (Cohen’s *d* = −0.41, pFDR = 0.049).

**Fig. 3.**
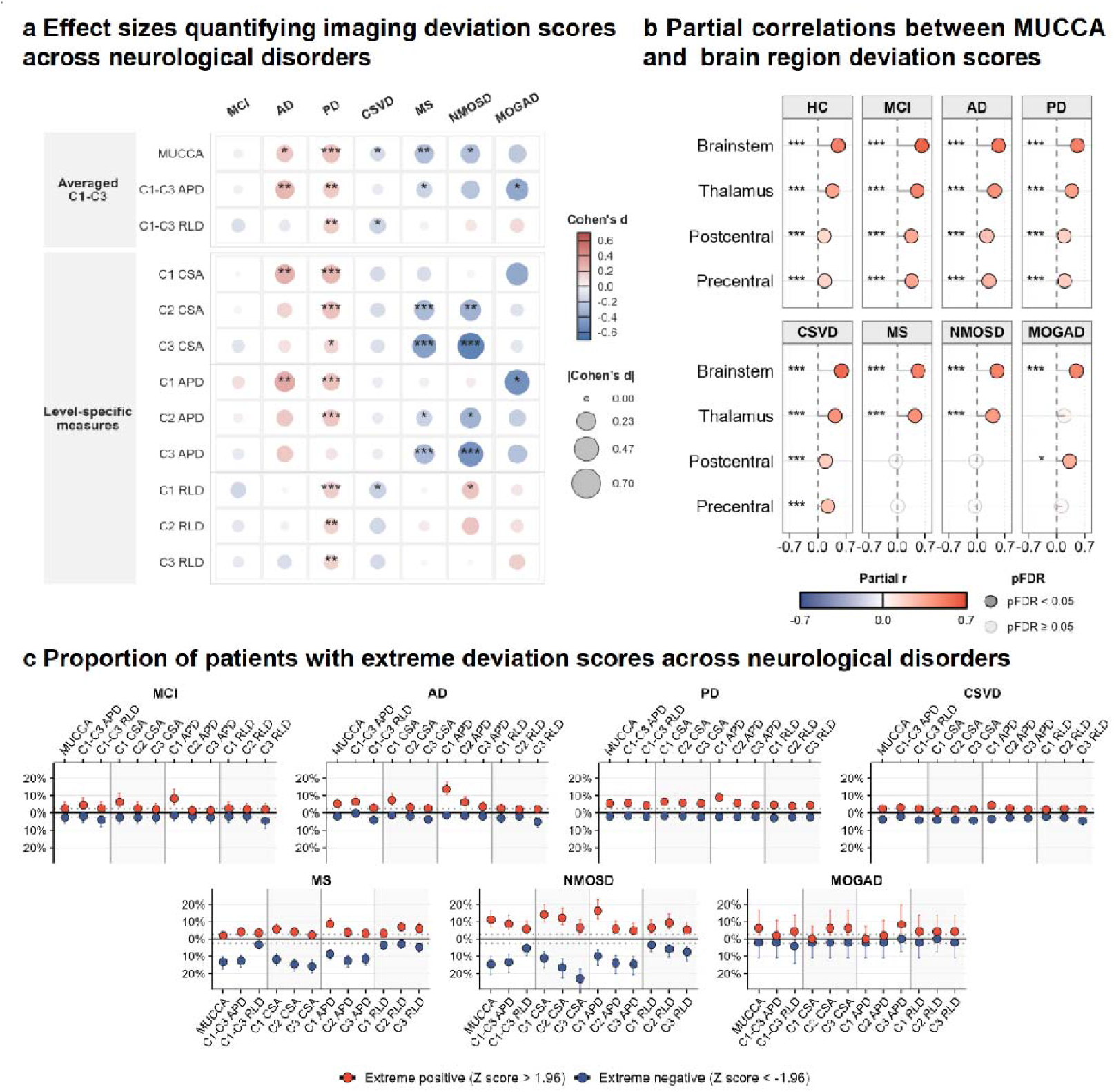
Heterogeneity in upper cervical spinal cord morphometric measures across neurological disorders. (a) The heatmap shows Cohen’s *d* effect sizes for group comparisons of upper cervical spinal cord Z scores between each disease group and HCs. Positive Cohen’s *d* values reflect increased Z scores compared to HCs. Darker colors represent larger effect sizes. Statistical significance was determined using Monte Carlo permutation tests with FDR correction (*, **, *** denote pFDR < 0.05, 0.01, 0.001, respectively). (b) Partial correlation between upper cervical spinal cord structure (MUCCA) deviation and regional brain deviations across both HCs and neurological disorders. Significance was determined by two-sided Pearson partial correlation analysis, controlling for age and sex with FDR correction (*, **, *** for pFDR < 0.05, 0.01, 0.001). (c) Proportions of extreme deviations in upper cervical spinal cord morphometric measures across neurological disorders. Dots indicate the proportions of extreme negative (< −1.96) and positive Z scores (> 1.96), and lines represent the corresponding 95% CIs. Note: MUCCA, mean upper cervical cord area; CSA, cross sectional area; APD, antero-posterior diameter; RLD, right-left diameter. HCs, healthy controls; MCI, mild cognitive impairment; AD, Alzheimer’s disease; PD, Parkinson’s disease; CSVD, cerebral small vessel disease; MS, multiple sclerosis; NMOSD, neuromyelitis optica spectrum disorder; MOGAD, myelin oligodendrocyte glycoprotein antibody-associated disease; ns, non-significant; CI, confidence interval; FDR, false discovery rate.

Level-specific analyses further revealed level-dependent deviation patterns that were not fully captured by averaged C1-C3 measures. Among neurodegenerative disorders, MCI showed no significant deviations at any upper cervical spinal cord level (C1, C2, or C3; all pFDR > 0.05). In AD, higher values were confined to the C1 level, including C1 CSA (Cohen’s *d* = 0.30, pFDR = 0.001) and C1 APD (Cohen’s *d* = 0.32, pFDR = 0.002). PD showed more widespread higher values across most measures (Cohen’s *d* = 0.14 to 0.27, all pFDR < 0.01) except for C3 APD. In PD, effect sizes were largest at C1 and progressively decreased from C1 to C3 for both CSA and APD. Vascular and inflammatory demyelinating diseases showed selective reductions in level-specific measures. CSVD showed lower values only in C1 RLD (Cohen’s *d* = −0.19, pFDR = 0.028); MS and NMOSD exhibited a similar reduction pattern, characterized by lower values in C2 CSA, C3 CSA, C2 APD, and C3 APD (MS: Cohen’s *d* = −0.45 to −0.19, all pFDR < 0.05; NMOSD: Cohen’s *d* = −0.61 to −0.35, all pFDR < 0.05), with larger absolute effect sizes at C3 than at C2 for CSA and APD. MOGAD showed a selective reduction only in C1 APD (Cohen’s *d* = −0.53, pFDR = 0.033).

To characterize extreme deviation patterns, individual deviation scores for each measure were classified into three categories: extreme negative deviation (Z score < −1.96), within the normative range (−1.96 ≤ Z score ≤ 1.96), and extreme positive deviation (Z score > 1.96). The proportions across all disease groups and morphometric measures are illustrated in **Fig. 3c**. Among the averaged C1-C3 measures, MS and NMOSD showed comparatively high proportions of extreme negative deviations in MUCCA (13.4% and 14.6%, respectively) and mean C1-C3 APD (12.6% and 13.5%, respectively), whereas extreme positive deviations were less prominent. Similar patterns were observed for level-specific measures, where extreme negative deviations in MS and NMOSD mainly involved level-specific CSA and APD measures. Additionally, AD and NMOSD showed higher proportions of extreme positive deviations for C1 APD (13.9% and 16.4%, respectively).

### Correlations between upper cervical spinal cord and brain structural deviations

Using MUCCA deviation scores as a representative measure of upper cervical spinal cord structure, we examined their associations with deviation scores of brain measures, including major brain morphometric measures (GMV, sGMV, WMV, CSF volume, cortical thickness, surface area, and cerebellum volume), as well as selected regional brain structures involved in sensorimotor pathways (brainstem, thalamus, postcentral gyrus, and precentral gyrus). Pearson correlation analyses with FDR correction were conducted with adjustment for age and sex. Across the major brain morphometric measures, MUCCA deviations were positively correlated with GMV, sGMV, WMV, CSF volume, cortical thickness, and surface area in most disease groups, while these associations were not observed in NMOSD (**Fig. S2b**). Across the selected regional brain structures involved in sensorimotor pathways, MUCCA deviations in HCs were positively correlated with deviations in all four brain structures, with the strongest correlation for the brainstem (r = 0.507, pFDR < 0.001), followed by the thalamus (r = 0.371, pFDR < 0.001). Similar patterns were observed in MCI, AD, PD, and CSVD. However, inflammatory demyelinating diseases exhibited a regionally selective MUCCA association: brainstem and thalamus in MS and NMOSD, brainstem and postcentral gyrus in MOGAD (**Fig. 3b**). To directly assess whether the strength of these associations differed from that in HCs, Fisher z-transformed correlation coefficients were compared between each disease group and HCs, with positive Fisher z differences (Δ z = z _HC_ – z _Disease_) indicating attenuated associations in the disease group relative to HCs. MS and NMOSD showed positive Fisher Δ z for the postcentral gyrus (Δ z = 0.188 to 0.191, both pFDR < 0.05) and precentral gyrus (Δ z = 0.169 to 0.225, both pFDR < 0.05) (**Fig. S2c**).

Disease-specific multivariable linear regression models were further fitted for each sensorimotor-related brain structure to assess whether the associations between MUCCA and brain deviations (brainstem, thalamus, postcentral gyrus, and precentral gyrus) varied with disease duration. Each model included an interaction term (MUCCA deviation × disease duration) and was adjusted for age and sex. Only the interaction for the thalamus in MOGAD remained significant, indicating that the association between MUCCA deviations and thalamic deviations changed with disease duration (interaction β = −1.546, 95% confidence interval [CI]: −2.474 to −0.619, pFDR = 0.018, **Fig. S2d**)

### Disease classification using upper cervical spinal cord deviations

To evaluate the stand-alone and incremental diagnostic value of upper cervical spinal cord deviations for disease classification, we established three Elastic Net-based models (α = 0.5) for a series of classification tasks: (1) Model 1: a baseline model including demographics (age and sex) and brain structural deviations; (2) Model 2: a spinal cord model including only upper cervical spinal cord deviations; and (3) Model 3: a combined model including demographics, brain structural deviations, and spinal cord deviations. Brain structural deviations included GMV, sGMV, WMV, CSF volume, cortical thickness, surface area, cerebellar volume, and intracranial volume (ICV). Classification tasks included disease versus HCs (MCI was not considered in these analyses, as it showed no significant differences from HCs in all upper cervical spinal cord measures) and four preselected pairwise disease classifications (NMOSD vs MOGAD, MS vs MOGAD, MS vs NMOSD, and MCI vs AD). Model performance was evaluated using nested ten-fold cross-validation (CV), with inner-loop elastic-net tuning and out-of-fold prediction. Area under the receiver operating characteristic curve (AUC) and 95% CIs were estimated from the pooled out-of-fold predictions using DeLong’s method. The incremental value was assessed using ΔAUC, defined as the AUC of the combined model minus that of the baseline model for each classification task.

As shown in **Fig. 4a**, the spinal cord model (Model 2) showed moderate-to-good discriminatory ability in three tasks: NMOSD versus MOGAD (AUC = 0.858, 95% CI: 0.807-0.908), NMOSD versus HCs (AUC = 0.837, 95% CI: 0.795-0.879), and MS versus MOGAD (AUC = 0.827, 95% CI: 0.768-0.886), while exhibiting poor or negligible discrimination in the remaining classification tasks. To assess the incremental value of upper cervical spinal cord deviations, we next compared the AUCs of the baseline (Model 1) and combined models (Model 3), with the ΔAUC serving as the measure of incremental value. For disease versus HCs classification, the largest significant ΔAUC was observed for NMOSD versus HCs (combined model: AUC = 0.906, 95% CI 0.879–0.933; ΔAUC = 0.103, 95% CI: 0.071-0.133, pFDR < 0.001), followed by MS versus HCs (combined model: AUC = 0.829, 95% CI 0.802-0.856; ΔAUC = 0.038, 95% CI: 0.024-0.051, pFDR < 0.001). For pairwise disease classification, the combined model outperformed the baseline model in distinguishing MS from NMOSD (combined model: AUC = 0.787, 95% CI: 0.748-0.825; ΔAUC = 0.039, 95% CI: 0.021-0.058, pFDR < 0.001) and MS from MOGAD (combined model: AUC = 0.870, 95% CI: 0.823-0.917; ΔAUC = 0.040, 95% CI: 0.005-0.077, pFDR = 0.047).

**Fig. 4.**
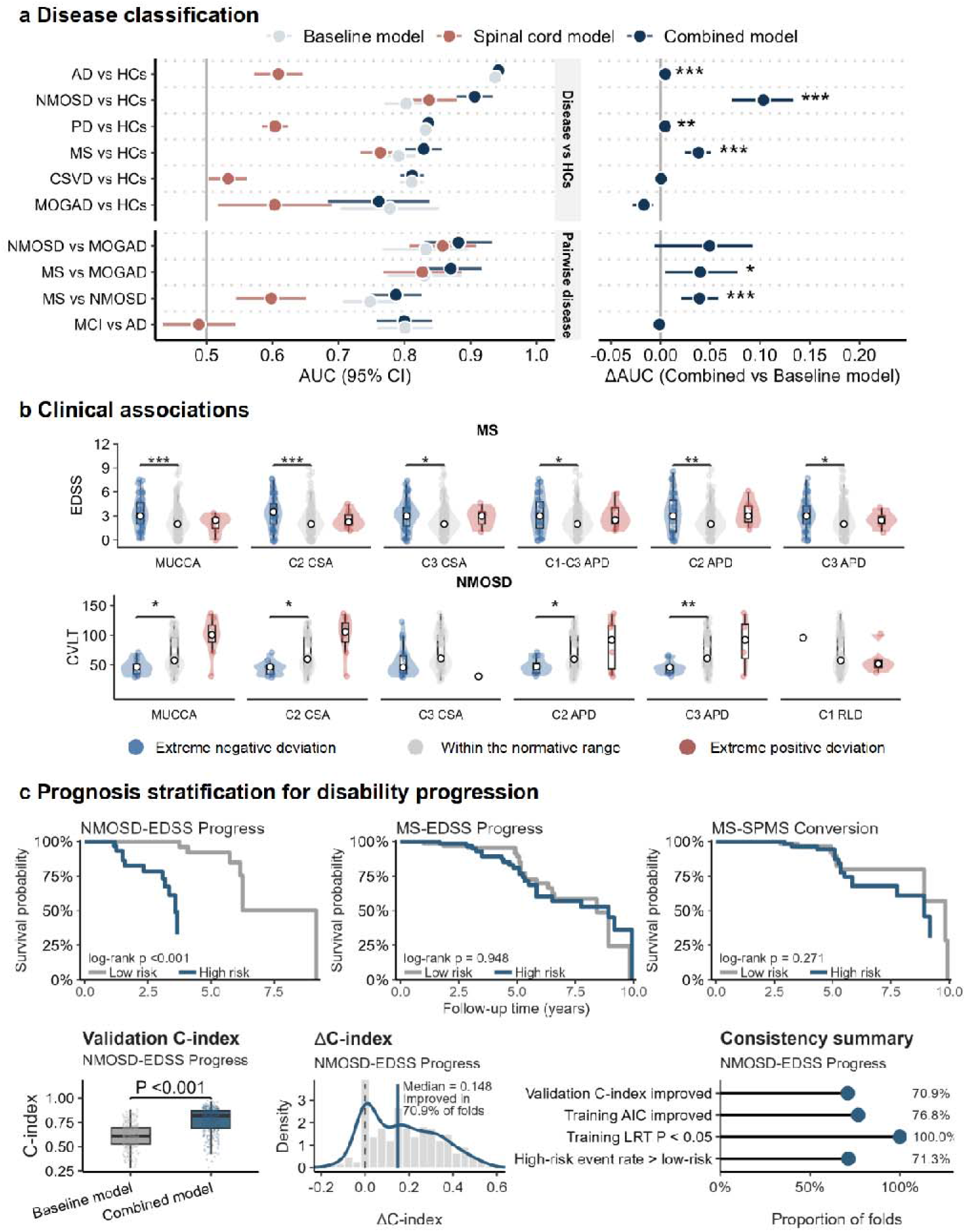
The value of upper cervical spinal cord deviations for disease differential diagnosis, clinical correlation, and prognosis evaluation. (a) Disease classification using upper cervical spinal cord deviations. Three models were evaluated: a baseline model incorporating age, sex, and brain structural deviations, a spinal cord model including upper cervical spinal cord deviations only, and a combined model incorporating age, sex, brain structural deviations and upper cervical spinal cord deviations. The left panel shows AUC (circle) and 95% CI (error bar) for each disease comparison. The right panel show ΔAUC (combined model vs baseline model) with 95% CI (error bar) for each disease comparison. Incremental diagnostic value was considered statistically significant when ΔAUC was > 0, the 95% CI excluded zero, and the FDR-adjusted p value was < 0.05. (*, **, *** for pFDR < 0.05, 0.01, 0.001). (b) Comparison of physical disability and cognitive performance among patients with extreme negative deviations (Z score < −1.96), values within the normative range (−1.96 to 1.96) and extreme positive deviations (Z score > 1.96) in MS and NMOSD. Pairwise group comparisons were performed using GLMs, adjusting for age and sex with FDR correction. *, **, *** for pFDR < 0.05, 0.01, 0.001. (c) Prognostic value of upper cervical spinal cord deviation scores in disability progression in NMOSD. The top row shows Kaplan-Meier curves stratified by the median predicted relative hazard from the combined model. The bottom row compares validation C-indices between the baseline (including age, sex, and baseline EDSS) and combined model (further including upper cervical spinal cord deviations), shows the distribution of ΔC-index across repeated cross-validation folds, and reports the proportion of folds with improved performance after adding spinal cord measures. Note: MUCCA, mean upper cervical cord area; CSA, cross sectional area; APD, antero-posterior diameter; RLD, right-left diameter; ICV, intracranial volume; AUC, area under the receiver operating characteristic curve; CI, confidence interval; HCs, healthy controls; AD, Alzheimer’s disease; PD, Parkinson’s disease; CSVD, cerebral small vessel disease; MS, multiple sclerosis; NMOSD, neuromyelitis optica spectrum disorder; MOGAD, myelin oligodendrocyte glycoprotein antibody-associated disease; SPMS, secondary progressive MS; FDR, false discovery rate; GLM, generalized linear models.

### Clinical associations with upper cervical spinal cord deviations

To assess the clinical relevance of deviation scores derived from normative modeling, between-group analyses were performed only for upper cervical spinal cord deviations that differed significantly between each disease group and HCs. Patients were stratified into three groups based on their deviation scores separately for each upper cervical spinal cord measure: extreme negative deviation (Z score < −1.96), within the normative range (−1.96 ≤ Z score ≤ 1.96), and extreme positive deviation (Z score > 1.96). MCI was not included in these analyses because it showed no significant differences from HCs across all upper cervical spinal cord measures, and MOGAD was excluded owing to its limited sample size. For the remaining disease groups, deviation-defined groups with 10 or fewer patients were excluded from the corresponding between-group analyses to minimize the impact of small sample sizes. Clinical characteristics, including physical disability, cognitive function, motor function, and CSVD burden, were compared across eligible deviation-defined groups using generalized linear models adjusted for age and sex. For each upper cervical spinal cord measure and disease group, FDR correction was applied for multiple comparisons across eligible deviation-defined groups.

As shown in **Fig. 4b**, regarding physical disability assessed by the Expanded Disability Status Scale (EDSS) score, patients with MS who showed extreme negative deviations in several upper cervical spinal cord measures, including MUCCA, C2 CSA, C3 CSA, mean C1-C3 APD, C2 APD, and C3 APD, had significantly higher EDSS scores than those within the normative range (all pFDR < 0.05). In patients with NMOSD, higher EDSS scores were observed only among those with extreme negative deviations in C3 CSA, compared with patients whose C3 CSA deviations were within the normative range (pFDR = 0.004, **Fig. S3c**). For cognitive performance, no significant between-group differences in Mini-Mental State Examination (MMSE) or Montreal Cognitive Assessment (MoCA) scores were observed in patients with AD, PD, CSVD, MS, or NMOSD (**Fig. S3a** and **Fig. S3b**). Additional cognitive assessments using the Brief Visuospatial Memory Test (BVMT) and California Verbal Learning Test (CVLT) were collected in patients with MS and NMOSD. In patients with NMOSD, those with extreme negative deviations in MUCCA, C2 CSA, C2 APD, and C3 APD had lower CVLT scores than those whose corresponding measures were within the normative range (all pFDR < 0.05, **Fig. 4b**). Similarly, patients with NMOSD who showed extreme negative deviations in C2 and C3 APD had lower BVMT scores than those with deviations within the normative range (all pFDR < 0.05, **Fig. S3c**). Regarding motor function in patients with PD, those with extreme positive deviations in C1 CSA had lower Part III of the Unified Parkinson’s Disease Rating Scale (UPDRS-III) scores (medication-off) than those with C1 CSA deviations within the normative range (pFDR = 0.041, **Fig. S3c**). CSVD burden, assessed using the CSVD score, did not differ significantly across the deviation-defined groups in patients with CSVD (all pFDR > 0.05, **Fig. S3c**).

### Prognostic value of upper cervical spinal cord deviations in disability progression

To assess the prognostic value of upper cervical spinal cord deviations for disability progression, we fitted Cox proportional hazards models for EDSS progression in MS and NMOSD, and for secondary progressive MS (SPMS) conversion in MS. MOGAD was not included in the prognostic modeling owing to the limited eligible sample size. Two Cox proportional hazards models were evaluated: a baseline model including age, sex, and baseline EDSS, and a combined model that further included upper cervical spinal cord deviations selected using elastic net Cox regression (α = 0.5). The median model-derived predicted relative hazard was used to stratify the MS and NMOSD patients into low- and high-risk groups. Kaplan-Meier analyses with log-rank tests revealed that NMOSD patients with a high predicted relative hazard had a greater risk of EDSS progression than those with a low predicted relative hazard (log-rank p < 0.001), whereas no significant difference between the low- and high-risk groups was observed for EDSS progression or SPMS conversion in MS (both log-rank p > 0.05, **Fig. 4c**).

Next, we assessed the incremental prognostic value of upper cervical spinal cord deviations for disability progression in MS and NMOSD patients, which was quantified using repeated stratified 3-fold internal CV with 100 repeats by comparing the C-index between the combined and the baseline models in the validation folds. Incremental performance was summarized by the change in C-index (ΔC-index) between the two models and the proportion of validation folds in which the combined model outperformed the baseline model. For EDSS progression in NMOSD, compared with the baseline model, the combined model showed improved prognostic discrimination. Specifically, the combined model achieved a higher validation C-index than the baseline model (p < 0.001). The ΔC-index distribution was shifted toward positive values, with a median gain of 0.148 and improvement observed in 70.9% of validation folds (**Fig. 4c**).

#### Clinically applicable personalized upper cervical spinal cord health report

To facilitate clinical translation, we developed a personalized health report. The report includes a summary of raw measures and deviation scores (Z scores), with extreme positive deviations (Z score > 1.96) and extreme negative deviations (Z score < −1.96) explicitly tagged (**Fig. 5**).

**Fig. 5.**
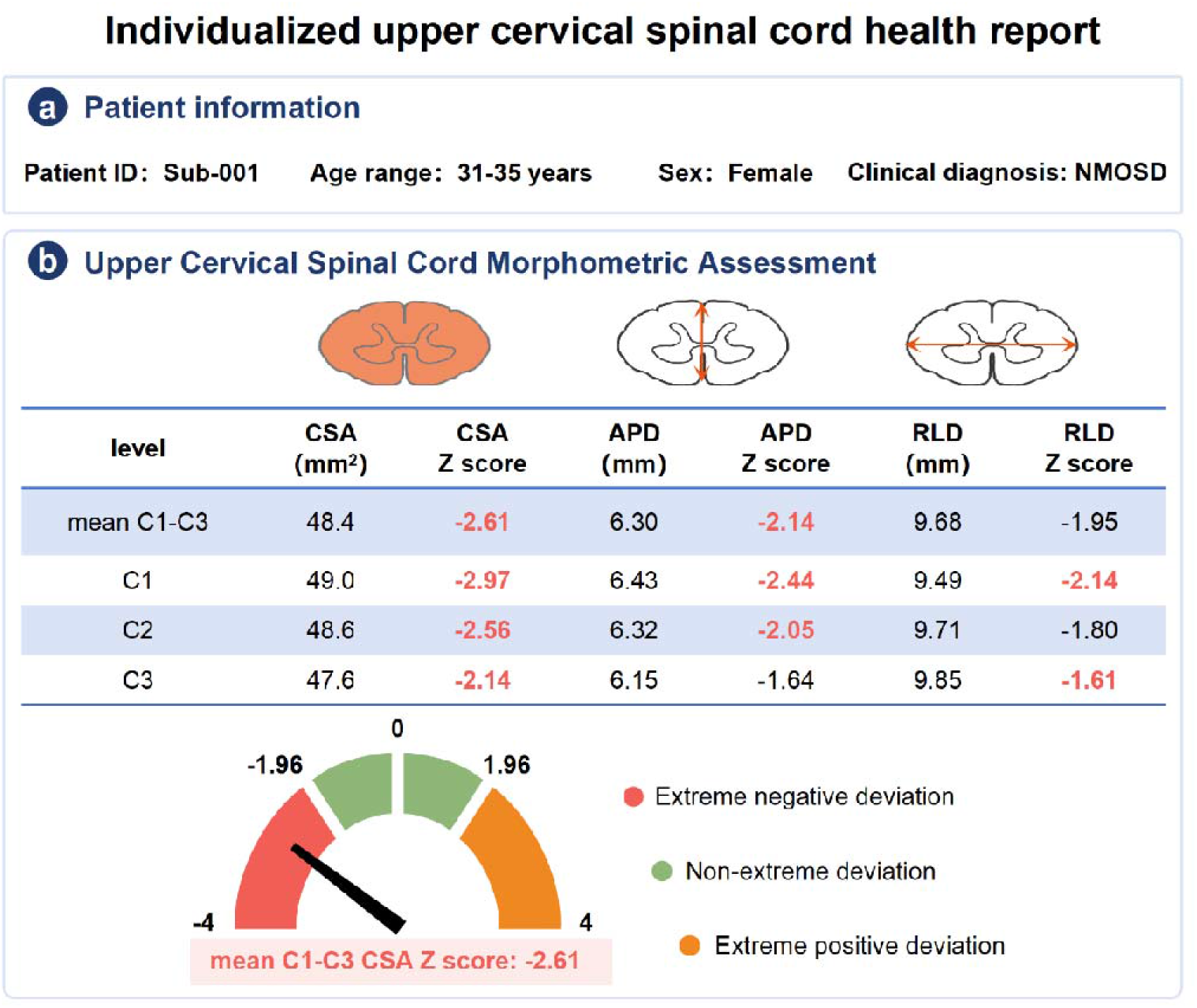
Personalized upper cervical spinal cord health reports generated from normative deviation scores. (a) Patient information. (b) Summary of the upper cervical spinal cord measures and deviation scores with extreme deviations explicitly flagged.

### Sensitivity analyses

We conducted sensitivity analyses to assess the robustness of our findings. Specifically, we examined the impact of ICV adjustment and age-range restriction on normative models, evaluated site effects using site-wise Z score distributions in HCs, refitted the diagnostic models and Cox proportional hazards models to include brain morphometric measures and sensorimotor-related regions, and compared deviation scores by upper cervical spinal cord lesion status in inflammatory demyelinating diseases.

First, ICV-adjusted normative models were established by including ICV as a fixed effect (**Fig. S4**). After ICV adjustment, all upper cervical spinal cord measures retained similar inverted U-shaped trajectories to those in the primary models. Agreement between the original and ICV-adjusted Z scores was assessed using the concordance correlation coefficient (CCC) and Bland-Altman analyses. CCC values ranged from 0.955 to 0.988 across all measures, and Bland-Altman analyses showed minimal bias between the original and ICV-adjusted Z scores, indicating that ICV adjustment had little effect on the derived normative deviation scores (**Fig. S5**).

Second, normative modeling was repeated in HCs aged 18-70 years, a more densely represented age range (**Fig. S6)**. Within this restricted age range, most upper cervical spinal cord measures showed relatively stable trajectories, with attenuated nonlinear patterns compared with the 8-85-year models. These findings suggest that younger (8-18 years) and older (70-85 years) age ranges contribute to the nonlinear lifespan trajectories observed in the primary analysis.

Third, site-wise Z score distributions among HCs were generally centered around zero for upper cervical spinal cord measures, with most site-specific medians close to the reference line (**Fig. S7**). Although variability was greater in some sites with smaller sample sizes, no systematic site-related shift was observed, supporting adequate site-wise calibration of the normative deviation scores.

Fourth, we refitted the diagnostic models for disease classification and Cox proportional hazards models for prognosis prediction by incorporating major brain morphometric measures and brainstem, thalamus, precentral gyrus, and postcentral gyrus. This analysis was performed to determine whether the incremental diagnostic value of upper cervical spinal cord deviations remained robust after additionally accounting for sensorimotor-related brain abnormalities. The incremental diagnostic value of upper cervical spinal cord deviations remained broadly consistent with the primary analysis, whereas their prognostic benefit was attenuated, with no consistent improvement in validation C-index and a median ΔC-index of approximately zero (**Fig. S8**).

Fifth, we conducted stratified analyses according to the presence or absence of MRI-visible cervical spinal cord lesions in patients with MS, NMOSD, and MOGAD. No significant differences in upper cervical spinal cord deviation scores were observed between patients with and without spinal cord lesions in any of the three diseases (all pFDR > 0.05, **Fig. S9**).

## Discussion

Using 3D T1w MRI data from 10,159 healthy individuals, we established age- and sex-specific normative references for upper cervical spinal cord morphology, including CSA, APD and RLD, both at the averaged C1-C3 level and individual cervical levels. All upper cervical spinal cord measures displayed an inverted U-shaped trajectory, with peak ages ranging from 29.7 to 44.1 years. Among these measures, CSA exhibited the most pronounced age-related change, whereas RLD followed a relatively flat trajectory. These trajectories closely aligned with those of white matter and brainstem volumes. Based on these normative references, we derived personalized deviation scores (Z scores) for HCs and patients, which captured disease-specific upper cervical spinal cord alterations and revealed disease-specific associations between upper cervical spinal cord and brain structures involved in sensorimotor pathways. Clinically, upper cervical spinal cord deviation scores facilitated disease classification, demonstrated significant associations with cognitive impairment and physical disability in MS and NMOSD, and predicted disability progression in NMOSD.

This study establishes a comprehensive normative framework for upper cervical spinal cord morphology across ages 8 to 85 years. By characterizing CSA, APD, and RLD at both the averaged C1-C3 level and individual cervical levels, we derived age-sensitive normative trajectories that enable individualized quantification of deviations in spinal cord morphology. All upper cervical spinal cord measures exhibited inverted U-shaped trajectories, with peak ages ranging from 29.7 to 44.1 years, a pattern partly supported by recent normative modeling studies^[16, 18]^. These trajectories provide age-sensitive references for distinguishing normal aging-related variation from disease-associated abnormalities^[12]^. We also observed that peak ages varied across morphometric dimensions and cervical levels, suggesting heterogeneous aging patterns of the upper cervical spinal cord. Relative rates of change further indicated that among the three morphometric dimensions, CSA was the most age-sensitive measure, while RLD showed comparatively modest age-related variation. After rescaling the normative trajectories of averaged upper cervical spinal cord morphology and brain structures to a standardized 0-1 range, averaged C1-C3 measures showed inverted U-shaped patterns similar to those of the brainstem and thalamus, which are anatomically plausible given their shared involvement in sensorimotor pathways^[20]^.

Beyond characterizing normative aging trajectories, recent work has begun to apply spinal cord normative modeling to clinical populations, including degenerative cervical disease^[17]^. We further applied this framework to neurological disorders. Based on the established normative models, averaged and level-specific analyses revealed both shared and disease-specific deviation patterns of upper cervical spinal cord morphology relative to HCs. For averaged C1-C3 measures, MCI showed no significant difference from HCs, which may indicate that upper cervical spinal cord involvement is limited or not yet detectable at this prodromal clinical stage. Other neurodegenerative diseases, including AD and PD, tended to show positive deviations in upper cervical spinal cord measures, although this observation should be interpreted in light of limited and partly inconsistent prior evidence^[5, 8]^. In contrast, CSVD and inflammatory demyelinating diseases, including MS, NMOSD, and MOGAD, showed negative deviation patterns, consistent with case-control studies reporting similar atrophy findings^[1, 2, 4]^. For level-specific measures, AD showed modest positive deviations restricted to the C1 level, including C1 CSA and C1 APD. This finding differs from previous small-sample case-control studies (both with fewer than 50 patients) that reported upper cervical spinal cord atrophy in AD compared with HCs^[5, 8]^. However, PD showed a modest but more widespread pattern, with positive effect sizes for CSA and APD being largest at the C1 level and progressively decreasing from C1 to C3 level. Although in vivo structural spinal cord studies in PD remain limited^[21]^, pathological evidence of observed axon swelling, which is associated with α-synuclein within the spinal cord of PD transgenic mice, may offer a pathological context for the positive deviations observed in PD^[22, 23]^. A recent normative modeling finding of positive brainstem and thalamic deviations in PD^[15]^ suggests that positive morphometric deviations in PD may not be confined to the upper cervical spinal cord. While AD and PD displayed a “rostral-to-caudal” pattern in level-specific analyses at C1-C3, MS and NMOSD followed a “caudal -to- rostral” pattern. In MS and NMOSD, negative deviations were observed in C2 CSA, C3 CSA, C2 APD, and C3 APD, with greater CSA and APD reductions at the C3 level than at the C2 level. This pattern may reflect spatially heterogeneous cervical cord atrophy in MS and NMOSD^[24, 25]^. These findings supported the complementary value of averaged and level-specific assessments in capturing disease-related upper cervical spinal alterations, while future work should further account for disease stage, treatment effect, and within-group heterogeneity.

Beyond characterizing disease-related deviation patterns in upper cervical spinal cord morphometry, we further examined whether the deviation scores of MUCCA, a representative measure of upper cervical spinal cord atrophy^[26]^, were associated with structural deviations in sensorimotor-related brain regions: the brainstem, thalamus, precentral gyrus, and postcentral gyrus. In HCs, individualized MUCCA deviation scores were significantly associated with deviation scores of these regional brain structures, consistent with the integrated functional organization of sensorimotor networks across the CNS^[27]^. Notably, in inflammatory demyelinating diseases (MS, NMOSD, and MOGAD), the associations between MUCCA deviation scores and sensorimotor-related brain regions showed a heterogeneous pattern, with some associations attenuated or absent and others preserved. The association between MUCCA and brainstem deviation scores was preserved across all three inflammatory demyelinating diseases. Together, these findings suggest that inflammatory demyelinating diseases may disrupt the normal structural association between the upper cervical spinal cord and sensorimotor-related brain structures. Longitudinal studies are needed to elucidate how lesion evolution, disease progression, relapse, and therapeutic interventions moderate brain-spinal cord structural associations.

We further evaluated the clinical utility of upper cervical spinal cord deviation scores in three clinical applications: (1) disease classification, (2) clinical association analyses, and (3) prognostic prediction. In disease classification, the spinal cord model incorporating upper cervical spinal cord deviations demonstrated moderate-to-good discriminatory ability in three classification tasks, including NMOSD versus MOGAD, NMOSD versus HCs, and MS versus MOGAD. Compared with the baseline model incorporating demographics and brain structural deviations, the combined model yielded incremental performance gains in several classification tasks. These findings support the value of upper cervical cord deviations for disease differentiation, suggesting that spinal cord deviation scores may capture disease-specific structural alterations that are not fully reflected by demographics and brain structural measures. For clinical association analyses, patients were stratified separately for each upper cervical spinal cord measure into groups with extreme negative deviations, values within the normative range, and extreme positive deviations. In these measure-specific analyses, extreme negative deviations in selected upper cervical spinal cord measures were associated with worse physical disability in MS and NMOSD and with poorer BVMT and CVLT performance in NMOSD. These findings suggest that pronounced negative deviations in specific upper cervical cord morphometric measures may reflect greater clinical severity in inflammatory demyelinating diseases, consistent with the known vulnerability of the spinal cord to inflammatory demyelination, axonal injury, and cumulative tissue loss in MS and NMOSD^[28, 29]^. However, these associations were not observed across all spinal cord measures, disease groups, or clinical domains, indicating that the clinical relevance of upper cervical cord deviations is likely measure- and disease-specific. For prognosis prediction, upper cervical spinal cord deviations added significant incremental value to basic clinical information for predicting EDSS progression in patients with NMOSD, although this contribution largely overlapped with that of brain measures. Finally, we translated this normative modeling framework into a clinically oriented upper cervical spinal cord report that summarizes individualized deviation scores and highlights the extent of morphometric deviation relative to age- and sex-specific normative references.

This study has several limitations. First, the retrospective multisite design may have introduced residual heterogeneity related to selection bias, protocol differences, scanner vendor, field strength, and acquisition parameters, despite adjustment for site effects in the normative models. Moreover, the absence of children younger than 8 years and sparse sampling in some age ranges may limit the generalizability and precision of the reference models. Second, the absence of subtype-level modeling may have masked subtype-specific profiles by pooling heterogeneous subgroups with potentially distinct patterns of upper cervical spinal cord involvement into group-level analyses. Third, using vertebral levels as anatomical references for spinal cord segment localization can introduce inter-individual variability, whereas alternative approaches based on anatomical landmarks such as the ponto-medullary junction^[30, 31]^ and spinal rootlets^[32, 33]^ enable more precise identification of spinal cord segments. Fourth, because this study was based on macrostructural morphometric measures derived from 3D T1w MRI, focal inflammatory lesion burden, including spinal cord T2 lesions, gadolinium-enhancing lesions, and cerebral white matter lesions, could not be quantified. Fifth, despite the internal CV in some analyses, the lack of longitudinal upper cervical spinal cord measures and independent external validation limits the generalizability of our findings. Prospective multicenter studies with balanced age distribution, subtype-specific analyses, multimodal imaging with lesion quantification, and independent external validation are warranted to improve the generalizability of the normative models and to clarify the potential biological basis of the observed deviations.

In conclusion, this study established normative references for upper cervical spinal cord morphometric measures using 3D T1w MRI data from 10,159 healthy individuals. Based on these references, we derived deviation scores that provide a refined, individualized framework for assessing upper cervical spinal cord abnormalities across multiple neurological disorders, characterizing disease-specific deviation patterns, facilitating disease classification, and linking these deviations to cognition and physical disability as well as prognostic outcomes. Furthermore, personalized upper cervical spinal cord reports could serve as a practical tool for routine clinical decision-making.

## Methods

### Study design

This multicenter neuroimaging study used a primarily cross-sectional design supplemented by longitudinal data, incorporating MRI and clinical data from 45 sites in China. First, we constructed normative references for upper cervical spinal cord morphometry using 3D T1w MRI data and derived individualized deviation scores (Z scores) for both HCs and patients with neurological disorders. We then examined associations between these deviation scores and sensorimotor-related brain structural deviations within each diagnostic group. The clinical utility of the deviation scores was evaluated in three tasks: (1) differential diagnosis using deviation-score-based models; (2) assessment of associations with clinical measures, including physical disability, cognitive function, motor function, and CSVD burden; and (3) prognostic prediction of disease progression in a longitudinal cohort. To assess the robustness of our findings, we performed additional sensitivity analyses examining the effects of ICV adjustment and age range restriction, and characterizing site-wise Z score distributions among HCs. In inflammatory demyelinating diseases, we further compared deviation scores between patients with and without MRI-visible upper cervical spinal cord lesions to assess the potential influence of focal lesions.

This study was approved by the Ethics Committees of Beijing Tiantan Hospital, Capital Medical University, Beijing, China (approval Nos. KY-2019-050-02, KY-2019-140-02, KY-2024-221-02 and KY-2025-203-03). Written informed consent was obtained from all participants or their legal guardians. The study adhered to the Strengthening the Reporting of Observational Studies in Epidemiology (STROBE) guidelines.

### Participants

This cross-sectional study included 10,159 HCs aged 8-85 years and 2,394 patients with one of the seven neurological disorders who underwent 3D T1w MRI with isotropic spatial voxel sizes of 1 mm or less. The patient groups comprised individuals with MCI (n = 157), AD (n = 309), PD (n = 911), CSVD (n = 416), MS (n = 382), AQP4+ NMOSD (n = 171), and MOGAD (n = 48).

Inclusion criteria were as follows: (1) the availability of high-quality 3D T1w MRI data; (2) HCs were defined as individuals between 8 and 85 years of age with no history of clinically diagnosed CNS disorders; (3) all patient diagnoses were made in accordance with internationally recognized clinical standards: MCI and AD were diagnosed according to the National Institute on Aging-Alzheimer’s Association (NIA-AA) criteria^[34]^; idiopathic PD was diagnosed according to the diagnostic criteria established by the International Parkinson and Movement Disorder Society^[35]^; CSVD was diagnosed based on the Standards for Reporting Vascular Changes on Neuroimaging (STRIVE) neuroimaging standards^[36]^; MS was diagnosed as relapsing-remitting MS based on the 2017 McDonald criteria, with confirmed seronegativity for both aquaporin-4 immunoglobulin G (AQP4-IgG) and myelin oligodendrocyte glycoprotein antibody (MOG-IgG)^[37]^; AQP4+ NMOSD was diagnosed according to the 2015 international consensus diagnostic criteria, with serum AQP4-IgG positivity detected by a cell-based assay^[38, 39]^; MOGAD was diagnosed according to the 2023 International MOGAD Panel proposed diagnostic criteria, with serum MOG-IgG positivity determined using a cell-based assay^[40]^.

Exclusion criteria were as follows: (1) incomplete MRI acquisition, significant image artifacts, or poor image quality as assessed by visual inspection; (2) a history of other neurological diseases, such as stroke or tumor; (3) failure of MR image segmentation or incomplete segmentation of the upper cervical spinal cord during image processing; (4) HCs and patients younger than 8 years or older than 85 years, owing to limited sample sizes in these age ranges.

### Demographic and clinical data collection

All participants were clinically evaluated, diagnosed, and monitored by neurologists with specialized expertise. Basic clinical information was collected, including age, sex, years of education, together with standardized assessment scales for physical disability, cognitive function, motor function, and CSVD burden. Physical disability was measured with the EDSS. Cognitive function was assessed across three domains. Global cognitive function was evaluated using the MMSE and MoCA. Verbal episodic memory was assessed with the CVLT, Third Edition ^[41]^, and visuospatial episodic memory with the BVMT-Revised^[42]^. Motor function in patients with PD was assessed using UPDRS-III in the medication-off state. For patients with CSVD, the CSVD score was used to reflect the overall MRI burden of the disease^[43]^. Extended follow-up assessments included EDSS progression in patients with MS and NMOSD, as well as conversion from relapsing-remitting MS to SPMS in patients with MS^[44]^.

### MRI data collection, segmentation and measure extraction

In this multicenter study, MRI data were acquired at hospitals across China using 1.5-T and 3.0-T scanners from different manufacturers, including Siemens Healthineers, GE HealthCare, Philips Healthcare, United Imaging Healthcare, and Neusoft Medical Systems (**Fig. S1e**). High-resolution 3D T1w images were obtained with an isotropic resolution of 1 mm or less, with available acquisition parameters summarized in **Table S1**. All MRI data underwent quality assessment by two independent reviewers (Z. Z., with 8 years of experience in MR post-processing, and Y. D., with 15 years of experience in neuroradiology), and discrepancies were adjudicated by a third senior neuroradiologist (Y. L., with 20 years of experience).

Upper cervical spinal cord segmentation was performed using SCT v6.0 (https://spinalcordtoolbox.com/)^[19]^. For each 3D T1w image, automatic spinal cord segmentation was performed using the deep learning-based sct_deepseg_sc algorithm^[45]^, followed by vertebral level identification using the sct_label_vertebrae algorithm^[46]^. All spinal cord segmentations and vertebral level labels were visually inspected for quality control, with representative examples shown in Fig. S10. Spinal cord morphometric measures were subsequently quantified, including three averaged C1-C3 measures (MUCCA, mean C1-C3 APD and RLD) and nine level-specific measures (CSA, APD, and RLD) at C1, C2, and C3.

Automated structural MRI segmentation of brain structural measures was conducted using FreeSurfer (version 7.3.2; http://surfer.nmr.mgh.harvard.edu). Standard preprocessing included non-uniformity correction, skull stripping, intensity normalization, and registration to Talairach space. The resulting brain segmentations were visually inspected and manually corrected when necessary. Segmentation quality was further assessed using the Euler number, an established FreeSurfer-based quality metric. Scans with Euler numbers more than two standard deviations below the median value across all sites were considered segmentation outliers^[9]^. We extracted a set of brain morphometric measures, including GMV, sGMV, WMV, CSF volume, mean cortical thickness, total cortical surface area, and total cerebellar volume, from the aseg.stats and aparc.stats outputs. In addition, brain structures involved in sensorimotor pathways, including the brainstem, thalamus, postcentral gyrus, and precentral gyrus, were selected as representative regions. These regions were chosen to capture key components of brainstem conduit, the thalamic integration center, the primary somatosensory cortex, and the primary motor cortex, respectively. Brainstem volume and bilateral thalamic volumes were extracted from the subcortical segmentation output (aseg.stats), and the cortical gray matter volumes of postcentral and precentral gyri were derived from the right and left cortical parcellation outputs (rh.aparc.stats and lh.aparc.stats). Bilateral thalamic and cortical regional volumes were summed across hemispheres for subsequent analyses.

### Normative modeling and deviation quantification

Normative models of upper cervical spinal cord morphometry were fitted using GAMLSS in R, based exclusively on data from eligible HCs (n = 10,159). Each spinal cord morphometric measure was modeled separately as the dependent variable. Prior to normative modeling, outliers were identified and excluded separately for each spinal cord measure among HCs using a mean ± 3 SD criterion, resulting in slight variation in modeling sample size across measures. The primary sex-stratified models incorporated age and sex as fixed-effect predictors. Site-related random-effect terms were evaluated to account for inter-site variability. Age effects were modeled using fractional polynomials in both the location and dispersion components. The GG distribution was used to accommodate skewed morphometric distributions^[12]^. Within this framework, the shape parameter (ν) was held constant, whereas the location (μ) and dispersion (σ) parameters were estimated. Candidate models with different fractional polynomial terms and random-effect terms were compared using the BIC, and the model with the lowest BIC was selected. Ten-fold CV was used to derive out-of-fold deviation scores for HCs, with each score generated from a normative model fitted without the corresponding participant.

Individualized deviation scores (Z scores) were derived for patients and HCs by mapping the observed value of each measure to its cumulative probability under the fitted, age- and sex-adjusted normative distribution, followed by inverse standard normal transformation^[9]^. For each upper cervical spinal cord measure, Z scores < −1.96 and > 1.96 indicated extreme negative and extreme positive deviations, respectively, whereas Z scores between −1.96 and 1.96 were considered to be within the normative range. These Z-score thresholds correspond to the lower and upper 2.5% of the normative distribution, respectively, and have been widely adopted in previous normative modeling studies^[13, 47, 48]^. For each spinal cord measure, peak age was defined as the age at which the model-derived median trajectory reached its maximum within the observed age range. To quantify uncertainty in peak age estimates, we performed bootstrap resampling with replacement (1,000 iterations) and refitted the model for each iteration. To characterize age-related morphometric changes in the upper cervical spinal cord, we calculated the relative rate of change (%/year) for each measure, defined as the first derivative of the fitted median trajectory divided by the corresponding predicted median value and multiplied by 100. The variability of these age-related changes was quantified using the SD of the relative rate of change across the given age range.

Additionally, to visualize the co-developmental and aging trajectories of upper cervical spinal cord and brain morphometric measures, normative models were also constructed for global and regional brain structural measures using the same GAMLSS framework. These models were obtained based on 3D T1w images from 28,650 healthy volunteers aged 8-85 years (median age, 42.00 years; IQR, 23.00-58.00 years).

### Statistical analyses

All statistical analyses were performed using R software (version 4.3.2). Normally distributed continuous variables are presented as mean ± SD, whereas non-normally distributed variables are presented as median and IQR. Categorical variables are presented as frequency and percentage [n (%)].

Upper cervical spinal cord structural metric deviation scores were compared between patients and HCs matched for age, sex, and site, where available. Group differences were assessed using Monte Carlo permutation tests with FDR correction. The healthy control group served as the reference population. Accordingly, Z scores for HCs were centered around 0. Positive Z scores indicated larger-than-expected structural measures, whereas negative Z scores indicated smaller-than-expected measures. Effect sizes, quantified using Cohen’s *d*, were used to quantify structural deviation from HCs, with positive values indicating higher upper cervical spinal cord measures relative to HCs.

To investigate the structural association between upper cervical spinal cord and brain deviations, age- and sex-adjusted partial Pearson correlation analyses were performed to examine associations between MUCCA deviations and deviations in brain structures involved in spinal cord-associated sensorimotor pathways. P values were corrected for multiple comparisons using the FDR method. To determine whether disease status altered the strength of these associations, Fisher z-transformed correlation coefficients were compared between each disease group and HCs, with positive Fisher z differences, defined as Δ z = z _HC_ - z _Disease_, indicating attenuated associations in the disease group relative to HCs. Furthermore, to assess whether associations between MUCCA deviations and selected brain structural deviations varied with disease duration, disease-specific interactions were examined using multivariable linear regression models for each brain region, with brain structural deviations as the outcome and MUCCA deviations, standardized disease duration, and their interaction (MUCCA deviation × disease duration) as predictors, adjusting for age and sex. FDR correction was applied to the interaction p values within each disease group across brain structures.

To assess the diagnostic value of upper cervical spinal cord deviations, we established three Elastic Net logistic regression models (α = 0.5) for each classification task: (1) a baseline model with age and sex forced in and brain structure deviations (WMV, GMV, sGMV, cerebellar volume, CSF volume, cortical thickness, surface area and ICV) selected by Elastic Net; (2) a spinal cord model including upper cervical spinal cord deviations alone, selected by Elastic Net; (3) a combined model with age and sex forced in, both brain structure deviations and spinal cord deviations selected by Elastic Net. Elastic Net logistic regression (α = 0.5) allowed automated feature selection and reduced potential multicollinearity among the spinal cord deviations. Model performance was evaluated using nested CV, with an outer ten-fold CV for out-of-fold prediction and model evaluation and an inner ten-fold CV for lambda (λ) optimization. The incremental diagnostic value of spinal cord deviations was assessed by comparing the AUC of the combined model with that of the baseline model using DeLong tests based on out-of-fold predicted probabilities. The 95% CIs for ΔAUC were estimated using 200 paired bootstrap iterations, and paired DeLong p values were adjusted for multiple comparisons using FDR correction. Incremental diagnostic value was considered statistically significant when ΔAUC was > 0, the 95% CI excluded zero, and the FDR-adjusted p value was < 0.05.

To explore clinical correlates of upper cervical spinal cord deviations, we compared clinical performance across subgroups stratified by deviation scores. For each upper cervical spinal cord measure, Z scores were classified as extreme negative (Z score < −1.96), within the normative range (−1.96 ≤ Z score ≤ 1.96), or extreme positive (Z score > 1.96). Deviation-defined groups containing 10 or fewer patients were excluded from the corresponding between-group analyses to reduce the potential influence of small sample sizes. Clinical characteristics, including physical disability, cognitive function, motor function, and CSVD burden, were compared across eligible deviation-defined groups using generalized linear models adjusted for age and sex. For each upper cervical spinal cord measure within each disease group, FDR correction was applied to account for multiple comparisons across eligible deviation-defined groups. Statistical significance was defined as pFDR < 0.05.

To assess the prognostic value of upper cervical spinal cord deviations for disability progression, we fitted Cox proportional hazards models for EDSS progression in MS and NMOSD, and for SPMS conversion in MS. MOGAD was not included in the prognostic modeling owing to the limited eligible sample size. Two Cox proportional hazards models were evaluated: a baseline model including age, sex, and baseline EDSS, and a combined model that further included upper cervical spinal cord deviations selected using elastic net Cox regression (α = 0.5). The median model-derived predicted relative hazard was used to stratify the MS and NMOSD patients into low- and high-risk groups. Kaplan-Meier curves were used to visualize model-derived risk stratification. Next, we assessed the incremental prognostic value of upper cervical spinal cord deviations for disability progression, which was quantified using repeated stratified 3-fold internal CV with 100 repeats by comparing the C-index between the combined and the baseline models in the validation folds. In each CV split, elastic net variable selection was performed within the training fold, and the held-out validation fold was used only for performance evaluation. Incremental performance was summarized by the change in C-index (ΔC-index) between the two models and the proportion of validation folds in which the combined model outperformed the baseline model.

### Sensitivity analyses

To evaluate the robustness of the normative modeling and downstream analyses, we conducted several sensitivity analyses. First, ICV-adjusted models were constructed by including ICV as a fixed effect, and their agreement with non-adjusted models was assessed using the CCC and Bland-Altman analysis. Second, normative modeling was repeated in the more densely represented age range (18-70 years) to assess the sensitivity of the findings to age-range restriction. Third, to assess potential residual site effects, site-wise Z score distributions among HCs were examined to determine whether normative deviation scores were centered around zero with comparable dispersion across sites. Fourth, we refitted the diagnostic models (for disease classification) and Cox proportional hazards models (for prognosis prediction) using brain structural deviations that also included sensorimotor-related brain regions. Fifth, in inflammatory demyelinating diseases, we compared deviation scores between patients with and without MRI-visible upper cervical spinal cord lesions to assess the potential influence of focal lesions.

## Acknowledgments

We thank all contributors to the Chinese Brain Health Imaging Consortium (CHART) for providing MRI data from healthy volunteers. This work was supported by the Key Special Project for Clinical Research of Tiantan Hospital (2025-A09) (YL), the Beijing Outstanding Young Scientist Program (JWZQ20240101025) (YL), the National Natural Science Foundation of China (NO. 82572171) (ZZ), Beijing Municipal Natural Science Foundation for Distinguished Young Scholars (No. JQ20035) (YL), Ministry of Science and Technology of China-National Key Research Project (2022YFC2009904) (YL), Beijing Hospital Management Center-Climb Plan (No. DFL20220503) (YL), Beijing Young Scholars (NO. 026) (YL), Capital Medical University Young Scholars (A2202) (YL), and Young Scientists Program of Beijing Tiantan Hospital, Capital Medical University (YSP202205) (ZZ).

## Data availability

All codes used for the statistical analyses and figures are publicly available at GitHub (https://github.com/JiajiaWang-TT/Normative-spinal-cord).

## Author contributions

Study design: Jiajia Wang, Yunyun Duan, Zhizheng Zhuo, Yaou Liu.

Data analysis: Jiajia Wang, Tiantian Hua, Tianyu Gao, Renlong Zhang.

Statistical analysis: Jiajia Wang, Zhizheng Zhuo, Rui Zhang, Yajun Yao.

Data collection: Jiajia Wang, Tiantian Hua, Rui Zhang, Siyao Xu, Cong Zhang, Li Chai, Tianyu Gao, Lu Bai, Bingjie Lin, Lifang Liao, Chao Zhang, Yuqi Luo, Xiaoqin Zhu, Demao Deng, Yuankui Wu, Haiyan Wu, Zhiqiang Zhang, Fuqing Zhou, Jiajia Zhu, Xi-Nian Zuo, Shufang Cheng, and Chinese Brain Health Imaging Consortium (CHART).

Manuscript writing: Jiajia Wang.

Manuscript editing: Cong Zhang, Lu Bai, Xiaozhou Du, Huiying Wang, Xi-Nian Zuo, Ya-Zhuo Kong, Julien Cohen-Adad, Ruomi Guo, Shuiwei Xia, Yunyun Duan, Zhizheng Zhuo, and Yaou Liu.

Other cntributors of Chinese Brain Health Imaging Consortium (CHART) who are not listed as authors are listed below.

Zhanjun Zhang^1^, Yaojing Chen^1^, Panjie Yan^1^, Mengjie Rong^1^, Liping Ai^2^, Xinxiang Zhao^2^, Feng Ao^3^, Yalin Bai^4^, Zheqi Gao^4^, Yutong Bai^5^, Jianguo Zhang^5^, Haihua Bao^6^, Ya Guo^6^, Qing Cai^7^, Chu-Chung Huang^7^, Jiancang Cao^8^, Lianping Zhao^8^, Feiyan Chen^9^, Hongjian He^9^, Feng Chen^10^, Fei Wang^10^, Kun Chen^11^, Zaixu Cui^12^, Yang Li^12^, Hui Dai^13^, Ningnannan Zhang^14^, Yuhui Du^15^, Guoguang Fan^16^, Ying Fu^17,18^, Lubin Gou^19,20^, Junqiang Lei^19,20^, Chunjie Guo^21^, Xuemei Han^22^, Yang Hou^23^, Basheng Hu^24^, Jianping Hu^25^, Peiyu Huang^26^, Xiaojun Xu^26^, Xiaojun Guan^26^, Hui-Jie Li^27^, Jianrui Li^28^, Junlin Li^29^, Songtao Yin^29^, Kexuan Li^30^, Zhenyu Shu^30^, Shaowu Li^31^, Lanxi Meng^31^, Wenling Li^32^, Li Zhu^32^, Xinyu Li^33^, Cheng Xu^33^, Yao Li^34^, Jie Luo^34^, Yongmei Li^35^, Yunfei Li^36^, Xiaohu Zhao^36^, Peipeng Liang^37^, Haiyan Liao^38^, Qin Shen^39^, Ching-Po Lin^39,40^, Jun Liu^41,42^, Jungang Liu^43^, Weiqi Liu^44^, Ying Liu^45^, Jun Qiu^45^, Shanshan Lu^46^, Feiyun Wu^46^, Su Lui^47^, Hailin Ma^48^, Ning Mao^49^, Linghui Meng^50^, Jian Zhao^50^, Yinfeng Qian^51^, Juwei Shao^52^, Decai Tian^53^, Yilong Wang^53^, Jun Xu^53^, Yiheng Tu^54^, Jinhui Wang^55,56,57^, Junkai Wang^58^, Wen Wang^59^, Xiaochun Wang^60^, Feifei Zhang^60^, Yong Wang^61^, Zheng Wang^62,63^, Jinyuan Weng^64^, Ying Wu^65^, Wei Wang^66^, Jiaheng Xu^67^, Tong Zhang^67^, Rui Xu^68^, Ruimeng Yang^69,70^, Xi Yin^71^, Yulong Yin^72^, Xin Zhao^72^, Rongjun Yu^73,74^, Zhen Yuan^75^, Hailian Zhang^76^, Yaqin Zhang^77^, Zhihua Zhang^78^, Xia Zhe^79^, Jiansong Ji^80^, Kuncheng Li^81^, Xujun Duan^82^.

Affiliations:

1 State Key Laboratory of Cognitive Neuroscience and Learning, Faculty of Psychology, Beijing Normal University, Beijing, China.

2 Department of Radiology, The Second Affiliated Hospital of Kunming Medical University, Kunming, China.

3 Department of Radiology, Renmin Hospital, Hubei University of Medicine, Shiyan, China.

4 Department of Radiology, Yuci Renmin Hospital, Jinzhong, China.

5 Department of Neurosurgery, Beijing Tiantan Hospital, Capital Medical University, Beijing, China.

6 Department of Medical Imaging Center, Qinghai University Affiliated Hospital, Xining, China.

7 School of Psychology and Cognitive Science, East China Normal University, Shanghai, China.

8 Department of Radiology, Gansu Provincial Hospital, Lanzhou, China.

9 School of Physics, Zhejiang University, Hangzhou, China.

10 Department of Radiology, Hainan General Hospital, Haikou, China.

11 Centre for Cognitive and Brain Sciences and Department of Psychology, University of Macau, Macau SAR, China.

12 Chinese Institute for Brain Research, Beijing, China.

13 Department of Radiology, Affiliated Hospital of Zunyi Medical University, Zunyi, China.

14 Department of Radiology and Tianjin Key Laboratory of Functional Imaging, Tianjin Medical University General Hospital, Tianjin, China.

15 School of Computer and Information Technology, Shanxi University, Taiyuan, China.

16 Department of Radiology, The First Hospital of China Medical University, Shenyang, China.

17 Department of Neurology and Institute of Neurology, The First Affiliated Hospital of Fujian Medical University, Fuzhou, China.

18 Institute of Neuroscience and Fujian Key Laboratory of Molecular Neurology, Fujian Medical University, Fuzhou, China.

19 Department of Radiology, Lanzhou University First Affiliated Hospital, Lanzhou, China.

20 Gansu Province Clinical Research Neurology, Center for Radiology Imaging, Lanzhou, China.

21 Department of Radiology, The First Hospital of Jilin University, Changchun, China.

22 Department of Neurology, China-Japan Union Hospital of Jilin University, Changchun, China.

23 Department of Radiology, Shengjing Hospital of China Medical University, Shenyang, China.

24 Department of Radiology, Nanfang Hospital, Southern Medical University, Guangzhou, China.

25 Department of Radiology, The First Affiliated Hospital of Fujian Medical University, Fuzhou, China.

26 Department of Radiology, The Second Affiliated Hospital of Zhejiang University School of Medicine, Hangzhou, China.

27 State Key Laboratory of Cognitive Science and Mental Health, Institute of Psychology, Chinese Academy of Sciences, Beijing, China.

28 Department of Diagnostic Radiology, Affiliated Jinling Hospital, Medical School of Nanjing University, Nanjing, China.

29 Department of Imaging Medicine, Inner Mongolia Autonomous Region People’s Hospital, Hohhot, China.

30 Department of Radiology, Zhejiang Province People’s Hospital, Hangzhou, China.

31 Beijing Neurosurgical Institute, Beijing, China.

32 Department of Radiology, General Hospital of Ningxia Medical University, Yinchuan, China.

33 Department of Imaging, Shanxi Provincial People’s Hospital, Taiyuan, China.

34 National Engineering Research Center of Advanced Magnetic Resonance Technologies for Diagnosis and Therapy, School of Biomedical Engineering, Shanghai Jiao Tong University, Shanghai, China.

35 Department of Radiology, The First Affiliated Hospital of Chongqing Medical University, Chongqing, China.

36 Department of Radiology, The Fifth People’s Hospital of Shanghai Fudan University, Shanghai, China.

37 School of Psychology, Capital Normal University, Beijing, China.

38 Department of Radiology, The Second Xiangya Hospital, Central South University, Changsha, China.

39 Institute of Neuroscience, National Yang Ming Chiao Tung University, Taipei, Taiwan.

40 Department of Education and Research, Taipei City Hospital, Taipei, Taiwan.

41 Department of Radiology, Chongqing University Central Hospital, Chongqing, China.

42 Department of Radiology, Chongqing Emergency Medical Center, Chongqing, China.

43 Department of Radiology, Xiamen Children’s Hospital (Children’s Hospital of Fudan University at Xiamen), Xiamen, China.

44 Sophmind Technology (Beijing), Beijing, China.

45 Department of Radiology, The First Affiliated Hospital of USTC, University of Science and Technology of China, Hefei, China.

46 Department of Radiology, The First Affiliated Hospital with Nanjing Medical University, Nanjing, China.

47 Huaxi MR Research Center (HMRRC), Department of Radiology, West China Hospital of Sichuan University, Chengdu, China.

48 Tibet Autonomous Region Key Laboratory for High Altitude Brain Science and Environmental Acclimatization, Lhasa, China.

49 Department of Radiology, Yantai Yuhuangding Hospital, Qingdao University, Yantai, China.

50 Department of Radiology, Hebei Medical University Third Hospital, Shijiazhuang, China.

51 Department of Radiology, The First Affiliated Hospital of Anhui Medical University, Hefei, China.

52 The Affiliated Hospital of Yunnan University, Kunming, China.

53 Department of Neurology, Beijing Tiantan Hospital, Capital Medical University, Beijing, China.

54 Department of Psychology, University of Chinese Academy of Sciences, Beijing, China.

55 Institute for Brain Research and Rehabilitation, South China Normal University, Guangzhou, China.

56 Key Laboratory of Brain, Cognition and Education Sciences (Ministry of Education), South China Normal University, Guangzhou, China.

57 Center for Studies of Psychological Application, South China Normal University, Guangzhou, China.

58 Department of Radiology, Aerospace Center Hospital, Beijing, China.

59 Department of Radiology and Functional and Molecular Imaging Key Lab of Shaanxi Province, Fourth Military Medical University, Xi’an, China.

60 Department of Radiology, First Hospital of Shanxi Medical University, Taiyuan, China.

61 Department of Radiology, The First Hospital of Hebei Medical University, Shijiazhuang, China.

62 School of Psychological and Cognitive Sciences, Peking University, Beijing, China.

63 School of Biomedical Engineering, Hainan University, Sanya, China.

64 Philips (China) Investment, Shanghai, China.

65 School of Medical Imaging, Shanxi Medical University, Taiyuan, China.

66 Department of Radiology, First Affiliated Hospital of Harbin Medical University, Harbin, China.

67 Department of Radiology, The Fourth Affiliated Hospital of Harbin Medical University, Harbin, China.

68 Department of Radiology, First Affiliated Hospital of Xinjiang Medical University, Urumqi, China.

69 Department of Radiology, The Second Affiliated Hospital, South China University of Technology, Guangzhou, China.

70 Department of Radiology, Guangzhou First People’s Hospital, Guangzhou, China.

71 Medical Imaging Center, The First Affiliated Hospital of Shihezi University, Shihezi, China.

72 School of Psychology, Northwest Normal University, Lanzhou, China.

73 Life Science Imaging Center, Hong Kong Baptist University, Hong Kong, China.

74 Department of Education and Psychology, Academy of Wellness and Human Development, Faculty of Arts and Social Sciences, Hong Kong Baptist University, Hong Kong, China.

75 Faculty of Health Sciences, University of Macau, Macau, China.

76 Department of Radiology, The Fifth People’s Hospital of Qinghai Province, Xining, China.

77 Department of Radiology, The Fifth Affiliated Hospital, Sun Yat-sen University, Zhuhai, China.

78 Department of Neurosurgery, Tsinghua University Yuquan Hospital (Tsinghua University Hospital of Integrated Traditional Chinese and Western Medicine), Beijing, China.

79 Department of MRI, Shaanxi Provincial People’s Hospital, Xi’an, China.

80 Zhejiang Key Laboratory of Imaging and Interventional Medicine, The Fifth Affiliated Hospital of Wenzhou Medical University, Lishui, China.

81 Department of Radiology, Xuanwu Hospital, Capital Medical University, Beijing, China.

82 The Clinical Hospital of Chengdu Brain Science Institute, School of Life Science and Technology, University of Electronic Science and Technology of China, Chengdu, China.

