## Supplementary information for "Upper cervical spinal cord morphometric charts: from population references to individualized clinical assessment"

### Supplementary Materials

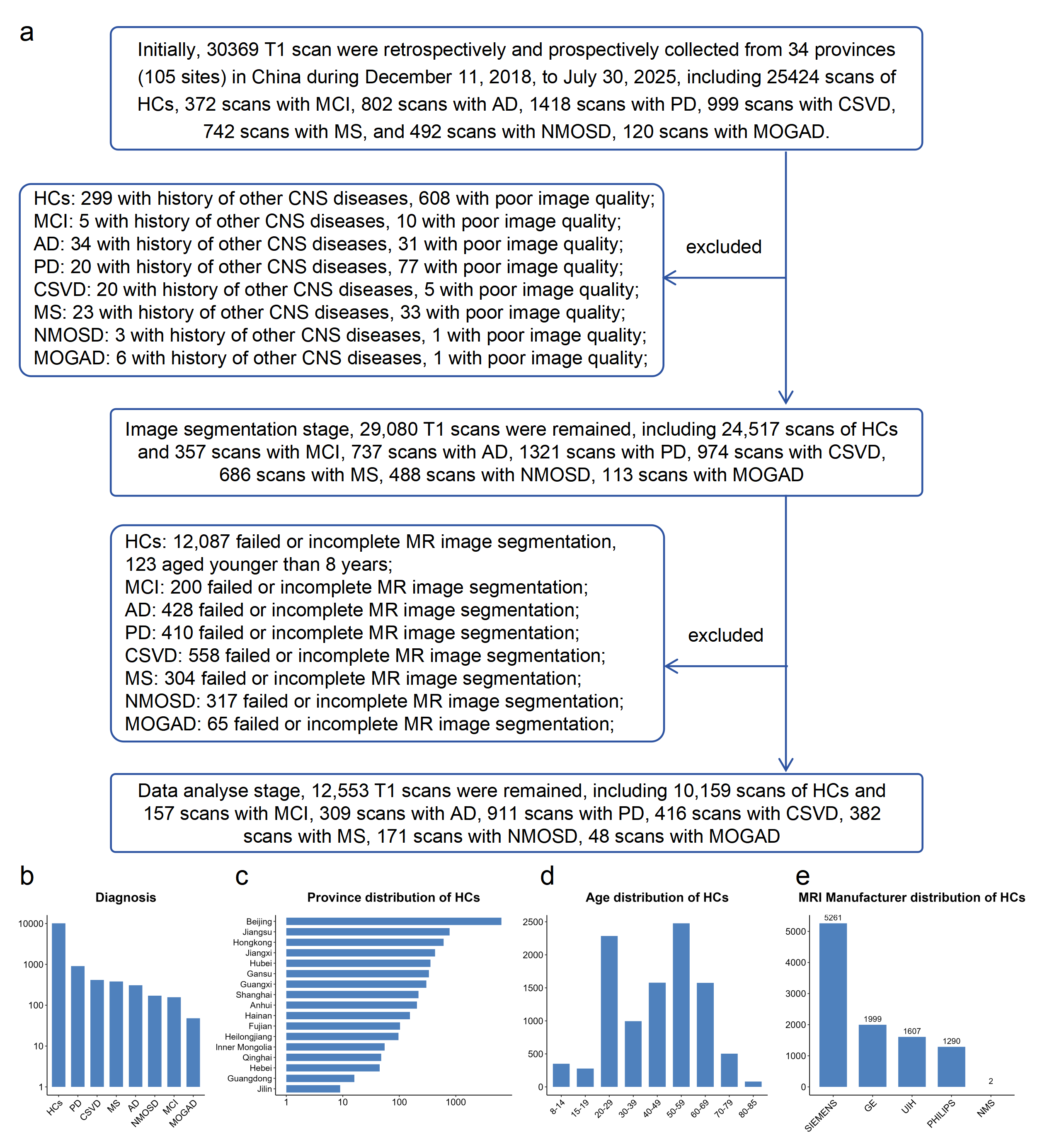
**Fig. S1. Overview of participant inclusion and demographic characteristics.** (a) Flowchart summarizing the inclusion process and final sample composition in this study. (b) Distribution of participants by diagnosis. (c) Distribution of HCs across provinces in China. (d) Age distribution of the included HCs. (e) Distribution of MRI scanner manufacturers among HCs.

Note: HCs, healthy controls; MCI, mild cognitive impairment; AD, Alzheimer’s disease; PD, Parkinson’s disease; CSVD, cerebral small vessel disease; MS, multiple sclerosis; NMOSD, neuromyelitis optica spectrum disorder; MOGAD, myelin oligodendrocyte glycoprotein antibody-associated disease; CNS, central nervous system; SIEMENS, Siemens Healthineers; GE, GE HealthCare; PHILIPS, Philips Healthcare; UIH, United Imaging Healthcare; NMS, Neusoft Medical Systems.

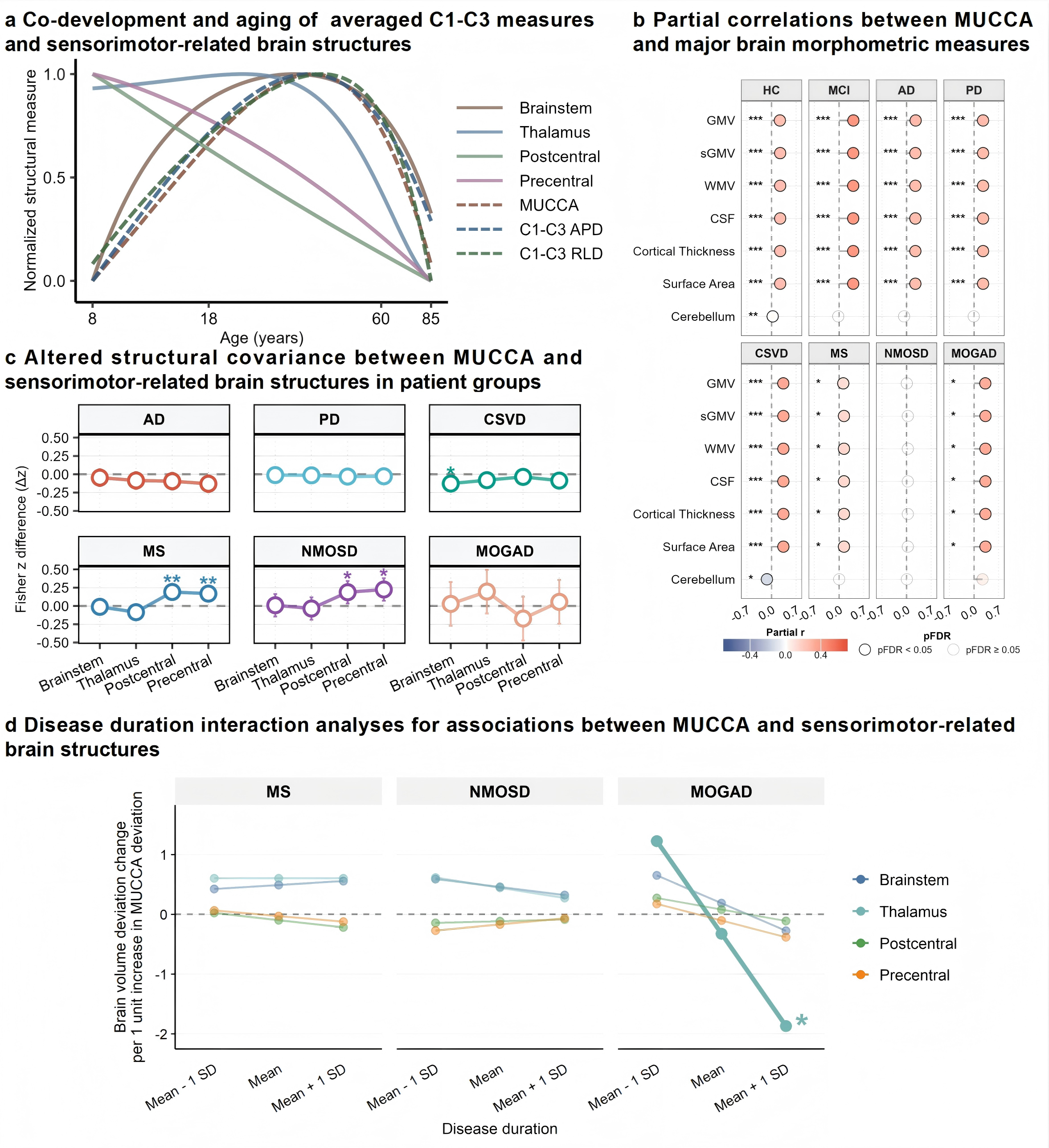
**Fig. S2. Patterns associated with age and disease in structural associations between the upper cervical spinal cord and sensorimotor-related brain structures.** (a) Normalized trajectories illustrating the co-development and aging patterns of averaged C1-C3 measures and sensorimotor-related brain structures. (b) Partial correlation between MUCCA deviation and major brain morphometric deviations across both HCs and neurological disorders. Significance was determined by two-sided Pearson partial correlation analysis, controlling for age and sex with FDR correction (*, **, *** for pFDR < 0.05, 0.01, 0.001). (c) Using HCs as the reference group, the decoupling index (Δ z = z _HC_ – z _Disease_) was calculated as the difference in Fisher z-transformed partial correlation of MUCCA with regional brain deviation scores between HCs and each disease group. The horizontal line at Δ z = 0 represents the coupling level between MUCCA and brain structures in HCs. Positive Δ z values indicate reduced coupling in patients relative to HCs. Dots and error bars represent Δ z values and 95% CIs, respectively. Asterisks denote regions that remained significant after within-disease FDR correction. (d) The moderating effect of disease duration on the association between MUCCA and four sensorimotor-related brain deviation scores in patients with MS, NMOSD, and MOGAD. Disease duration was modeled as a continuous variable and visualized at three representative levels: mean - 1 SD, mean, and mean + 1 SD. The y-axis shows the estimated change in regional brain volume deviation per 1-unit increase in MUCCA deviation. Values above and below zero indicate positive and negative spinal cord–brain associations, respectively. Thicker lines and asterisks denote statistically significant MUCCA × disease duration interactions after FDR correction. *, ** for pFDR < 0.05, 0.01.

Note: MUCCA, mean upper cervical cord area; CSA, cross‑sectional area; APD, antero-posterior diameter; RLD, right-left diameter; CI, confidence interval; HCs, healthy controls; SD, standard deviation.

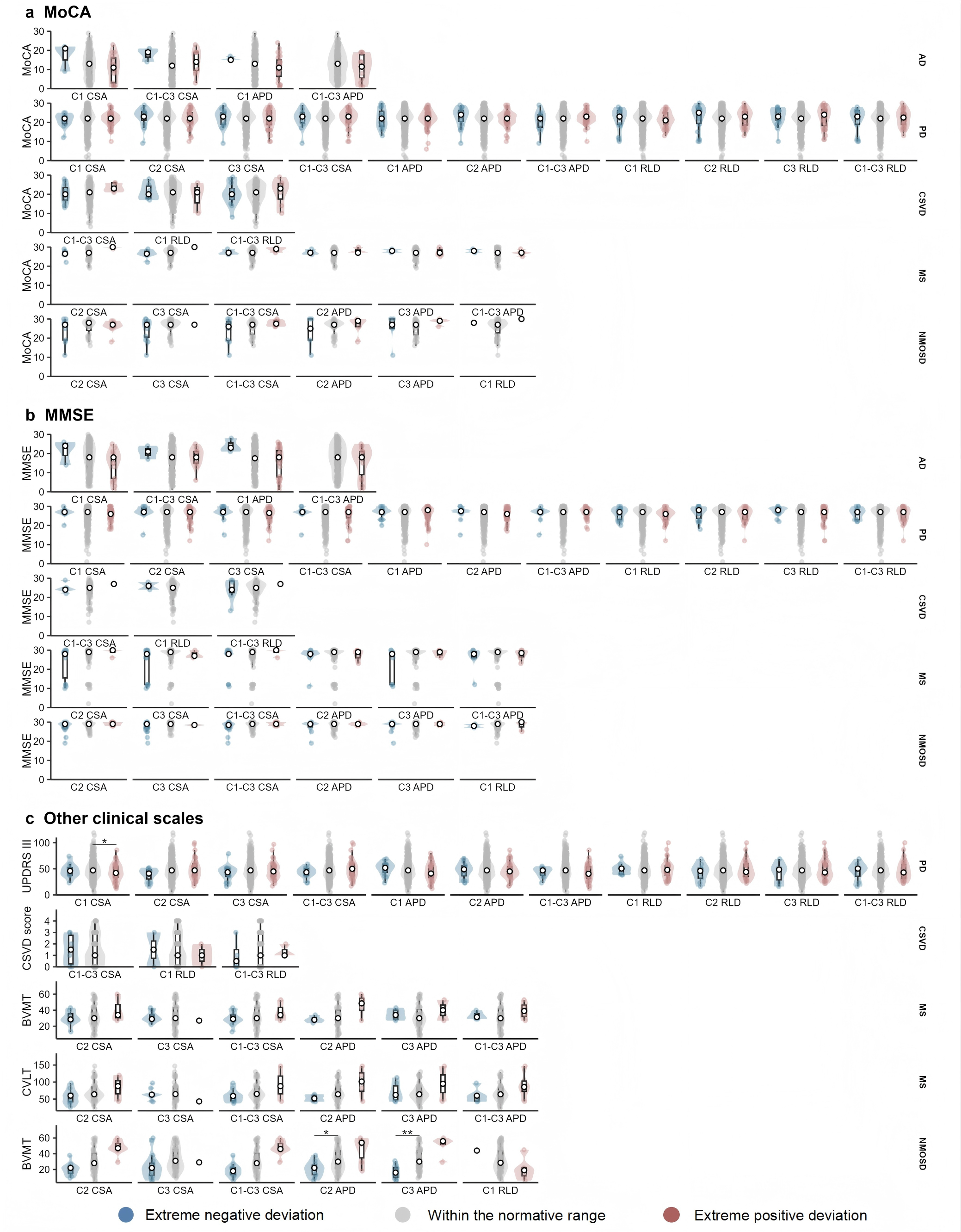

**Fig. S3. Comparison of clinical performance among patients with extreme negative deviations, values within the normative range, and extreme positive deviations in upper cervical spinal cord measures.** For each imaging measure within each disease group, participants were categorized based on the corresponding Z scores as extreme negative (Z score < −1.96), within the normative range (−1.96 ≤ Z score ≤ 1.96), and extreme positive (Z score > 1.96). Pairwise group comparisons were performed using GLMs, adjusting for age and sex with FDR correction. *, **, *** for pFDR < 0.05, 0.01, 0.001.

Note: MMSE, Mini-Mental State Examination; MoCA, Montreal Cognitive Assessment; GLM, generalized linear models; FDR, false discovery rate; CSA, cross-sectional area; APD, antero-posterior diameter; RLD, right-left diameter; UPDRS-III, Part III of the Unified Parkinson’s Disease Rating Scale; CVLT, California Verbal Learning Test; BVMT, Brief Visuospatial Memory Test.

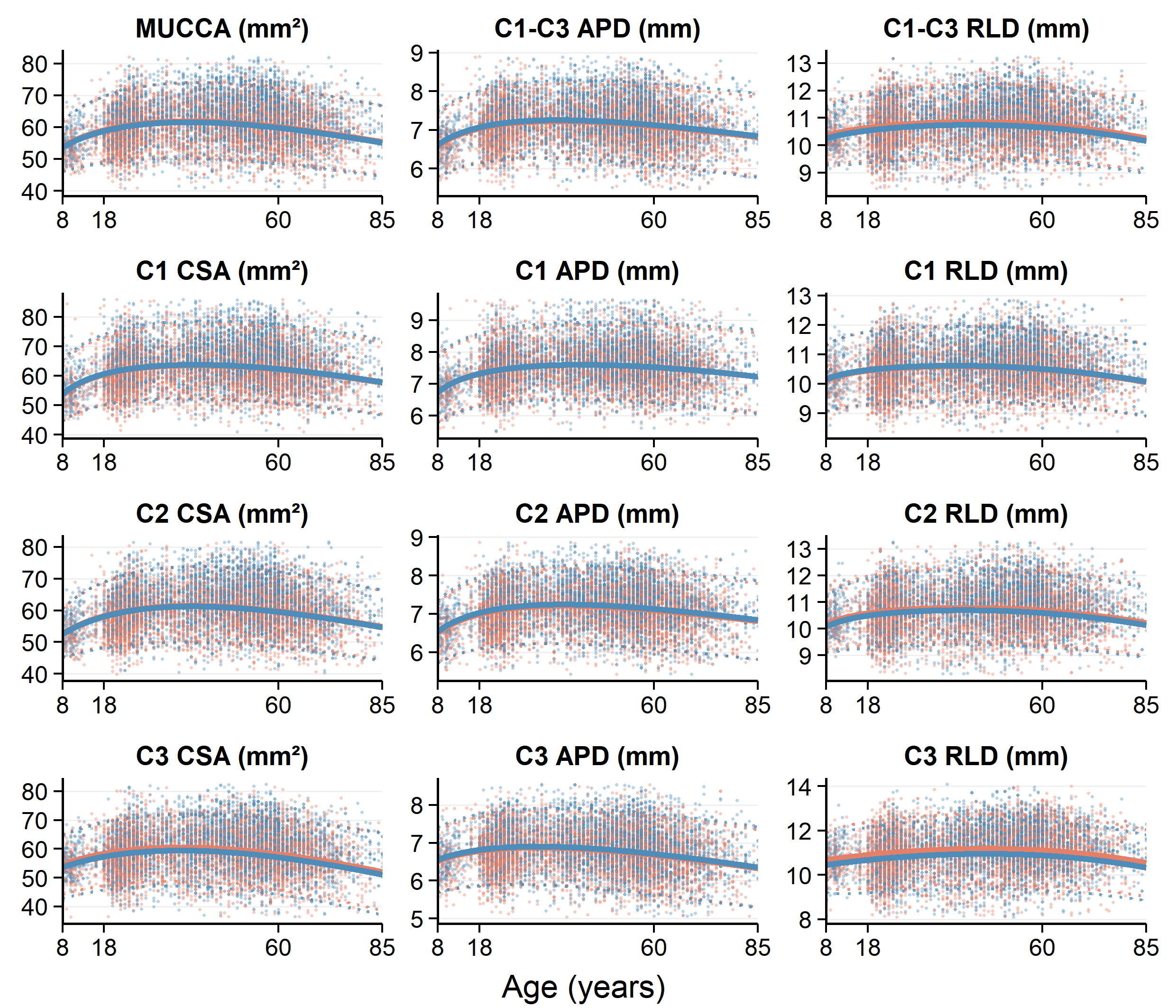

**Fig. S4. Normative trajectories of upper cervical spinal cord morphometric measures across ages 8-85 years after ICV adjustment.** Trajectories were estimated using GAMLSS, with age, sex, and ICV included as fixed effects, and site modeled as a random effect to account for inter-site variability. Solid lines indicate sex-specific medians, with shaded bands for the 95% CI.

Note: ICV, intracranial volume; GAMLSS, generalized additive models for location, scale, and shape; MUCCA, mean upper cervical cord area; CSA, cross‑sectional area; APD, antero-posterior diameter; RLD, right-left diameter; CI, confidence interval.

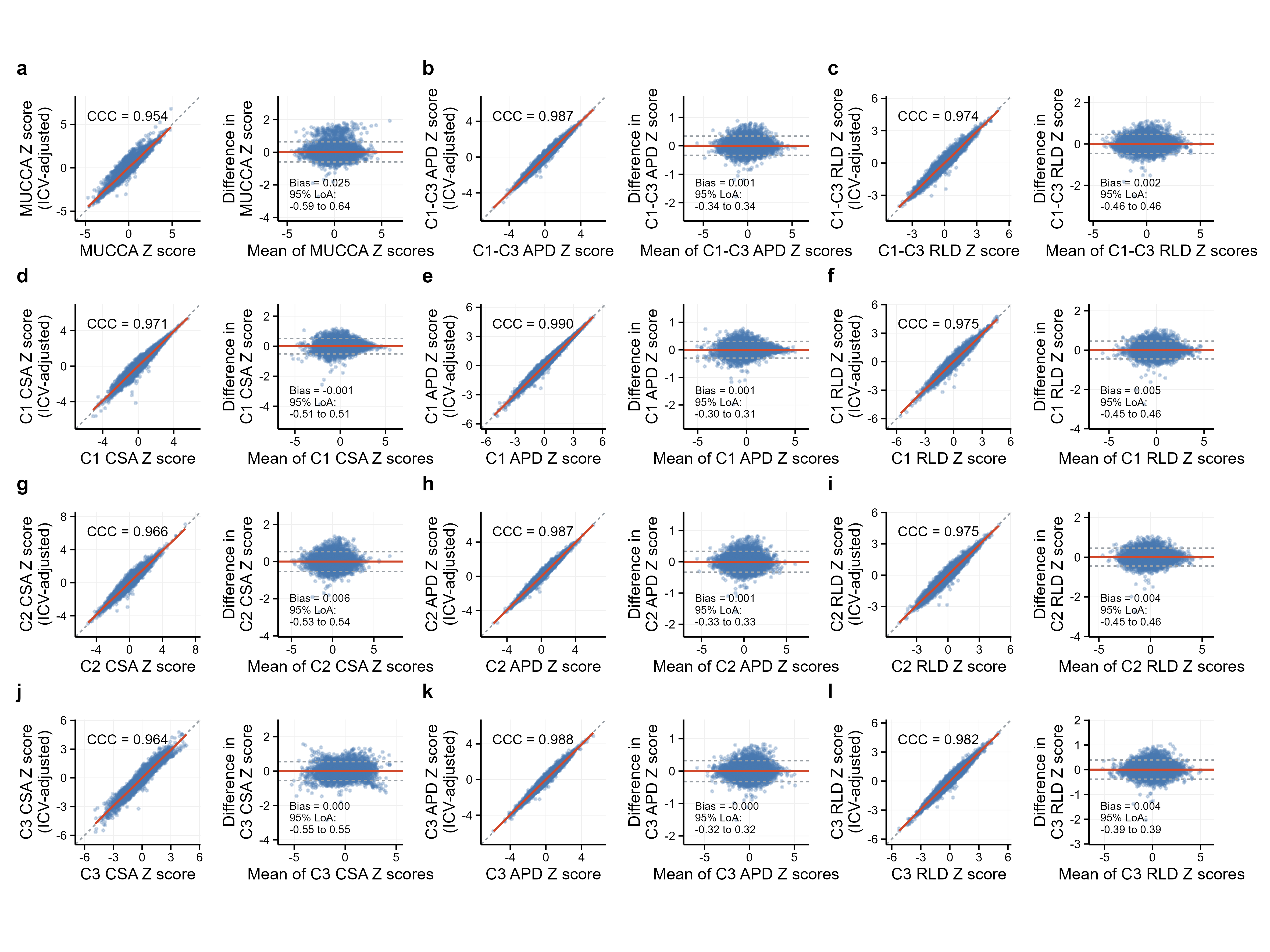
**Fig. S5. The agreement between original and ICV-adjusted Z scores for upper spinal cord measures.** Scatter plots and Bland-Altman plots compared the Z scores derived from the original normative model and the ICV-adjusted model across upper cervical spinal cord measures: MUCCA (a), mean C1-C3 APD (b), mean C1-C3 RLD (c), C1 CSA (d), C1 APD (e), C1 RLD (f), C2 CSA (g), C2 APD (h), C2 RLD (i), C3 CSA (j), C3 APD (k), C3 RLD (l). For each measure, the scatter plot (left panel) shows the concordance between original and ICV-adjusted Z scores, with the CCC displayed within the panel and the identity line shown as a gray dashed reference line. The corresponding Bland-Altman plot (right panel) displays the difference (ICV-adjusted Z score minus original Z score) against the mean of the two Z scores, with the red solid line indicating the bias (mean difference), gray dashed lines indicating the 95% limits of agreement (bias ± 1.96 SD).

Note: MUCCA, mean upper cervical cord area; CSA, cross‑sectional area; APD, antero-posterior diameter; RLD, right-left diameter; SD, standard deviation; ICV, intracranial volume; CCC, concordance correlation coefficient; LoA, limits of agreement.

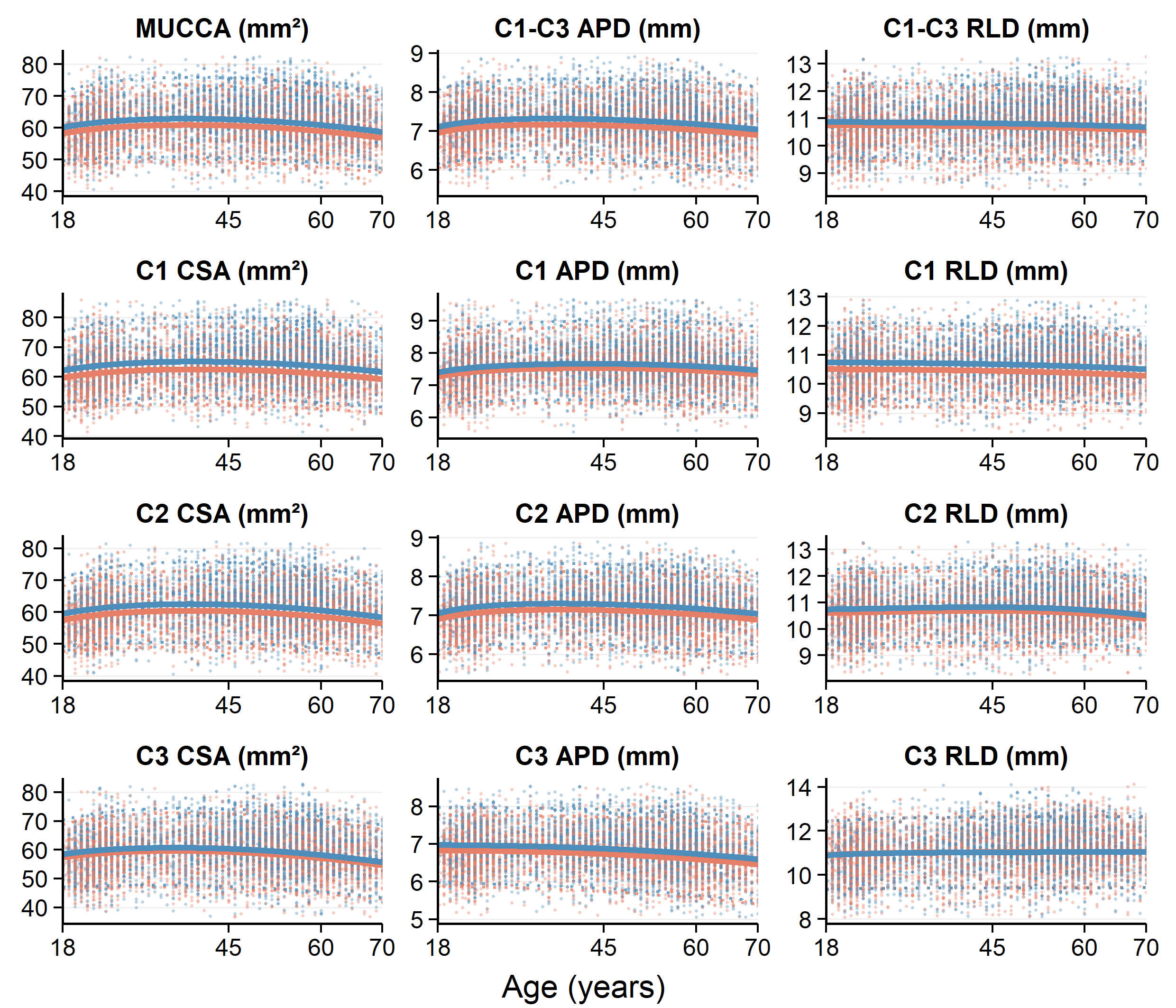

**Fig. S6. Normative trajectories of upper cervical spinal cord morphometric measures across ages 18-70 years.** Trajectories were estimated using GAMLSS, with age and sex included as fixed effects, and site modeled as a random effect to account for inter-site variability. Solid lines indicate sex-specific medians, with shaded bands for the 95% CI.

Note: GAMLSS, generalized additive models for location, scale, and shape; MUCCA, mean upper cervical cord area; CSA, cross‑sectional area; APD, antero-posterior diameter; RLD, right-left diameter; CI, confidence interval.

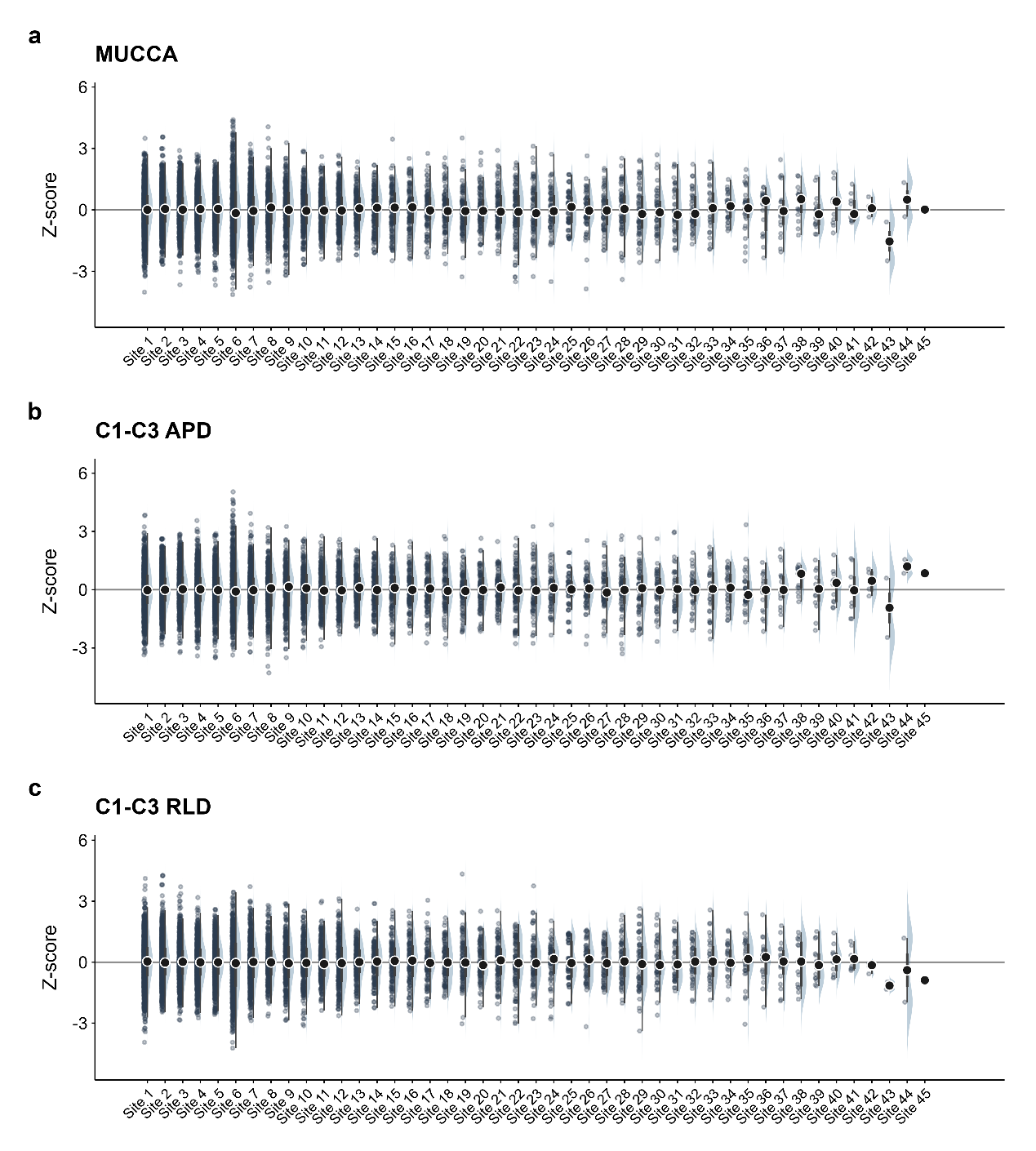
**Fig. S7.** Site-wise distributions of Z scores for averaged upper cervical spinal cord measures in HCs. Half-violin plots show the site-wise distributions of Z scores for MUCCA, mean C1-C3 APD, and mean C1-C3 RLD in HCs. Acquisition sites were anonymized as Site 1 to Site n and ordered in descending order according to the number of HCs at each site. Individual data points are shown on the left side of each half-violin plot. Black points and short horizontal bars indicate site-specific medians, and embedded boxplots show the IQR. The solid horizontal line indicates a Z score of 0. Overall, the distributions were centered around zero across most sites, supporting adequate site-wise calibration of the normative deviation scores.

Note: HCs, healthy controls; MUCCA, mean upper cervical cord area; APD, antero-posterior diameter; RLD, right-left diameter; IQR, interquartile range.

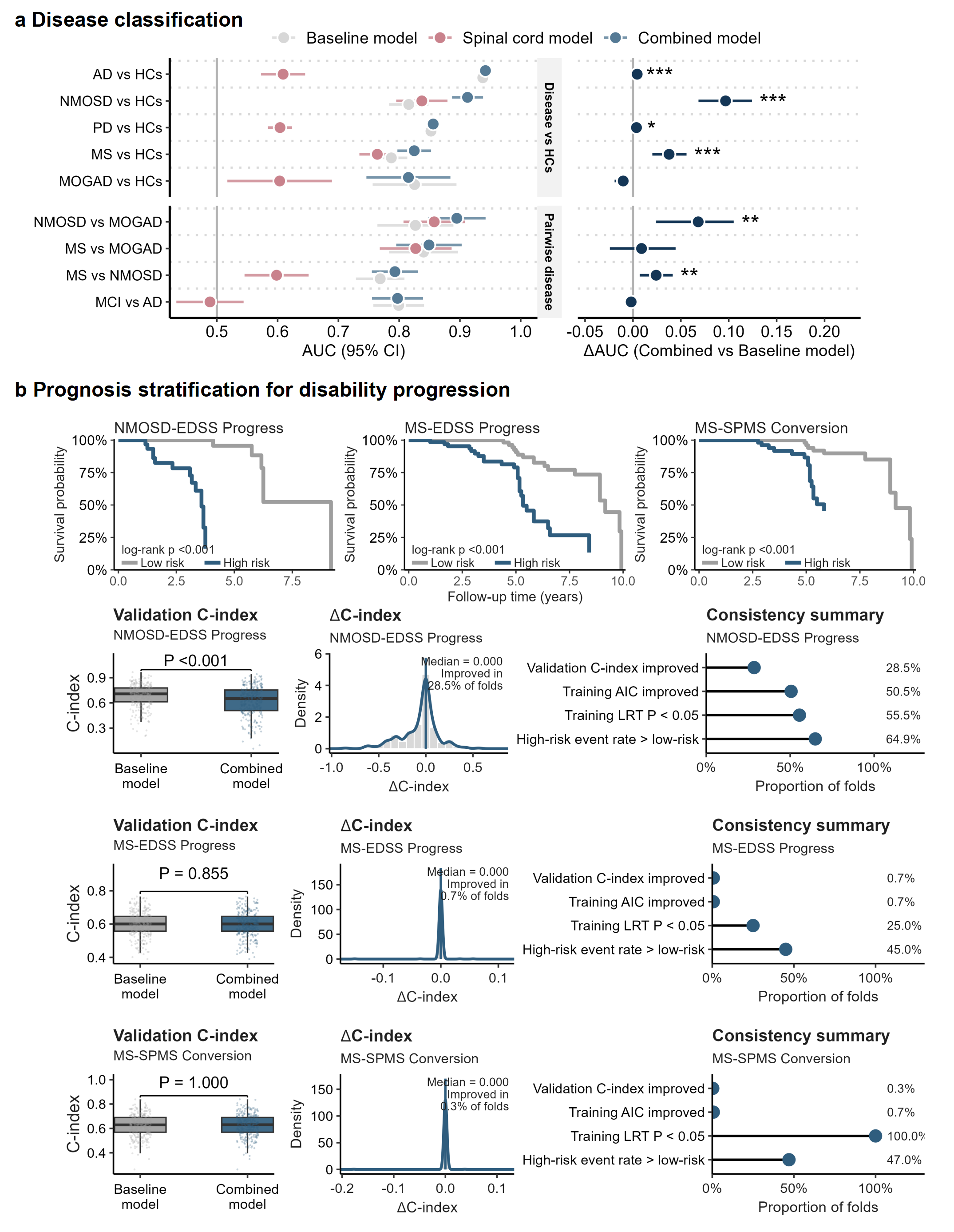
**Fig. S8.** The value of upper cervical spinal cord deviations for disease differential diagnosis and prognosis evaluation. (a) Disease classification using upper cervical spinal cord deviations. Three models were evaluated: a baseline model incorporating age, sex, and brain structural deviations, a spinal cord model including upper cervical spinal cord deviations only, and a combined model incorporating age, sex, brain structural deviations and upper cervical spinal cord deviations. Brain measures included WMV, GMV, sGMV, cerebellar volume, CSF volume, cortical thickness, surface area, ICV, brainstem, thalamus, postcentral gyrus, and precentral gyrus. The left panel shows AUC (circle) and 95% CI (error bar) for each disease comparison. The right panel show ΔAUC (combined model vs baseline model) with 95% CI (error bar) for each disease comparison. Incremental diagnostic value was considered statistically significant when ΔAUC was > 0, the 95% CI excluded zero, and the FDR-adjusted p value was < 0.05. (*, **, *** for pFDR < 0.05, 0.01, 0.001). (b) Prognostic value of upper cervical spinal cord deviation scores in disability progression in NMOSD. The top row shows Kaplan-Meier curves stratified by the median predicted relative hazard from the combined model. The second through fourth rows compare validation C-indices between the baseline (including age, sex, baseline EDSS, and brain structural deviations) and combined model (further including upper cervical spinal cord deviations), shows the distribution of ΔC-index across repeated cross-validation folds, and reports the proportion of folds with improved performance after adding spinal cord measures. Brain measures included WMV, GMV, sGMV, cerebellar volume, CSF, cortical thickness, surface area, ICV, brainstem, thalamus, postcentral gyrus, and precentral gyrus.

Note: MUCCA, mean upper cervical cord area; CSA, cross‑sectional area; APD, antero-posterior diameter; RLD, right-left diameter; ICV, intracranial volume; AUC, area under the receiver operating characteristic curve; CI, confidence interval; HCs, healthy controls; AD, Alzheimer’s disease; PD, Parkinson’s disease; CSVD, cerebral small vessel disease; MS, multiple sclerosis; NMOSD, neuromyelitis optica spectrum disorder; MOGAD, myelin oligodendrocyte glycoprotein antibody-associated disease; SPMS, secondary progressive MS; FDR, false discovery rate; GLM, generalized linear models.

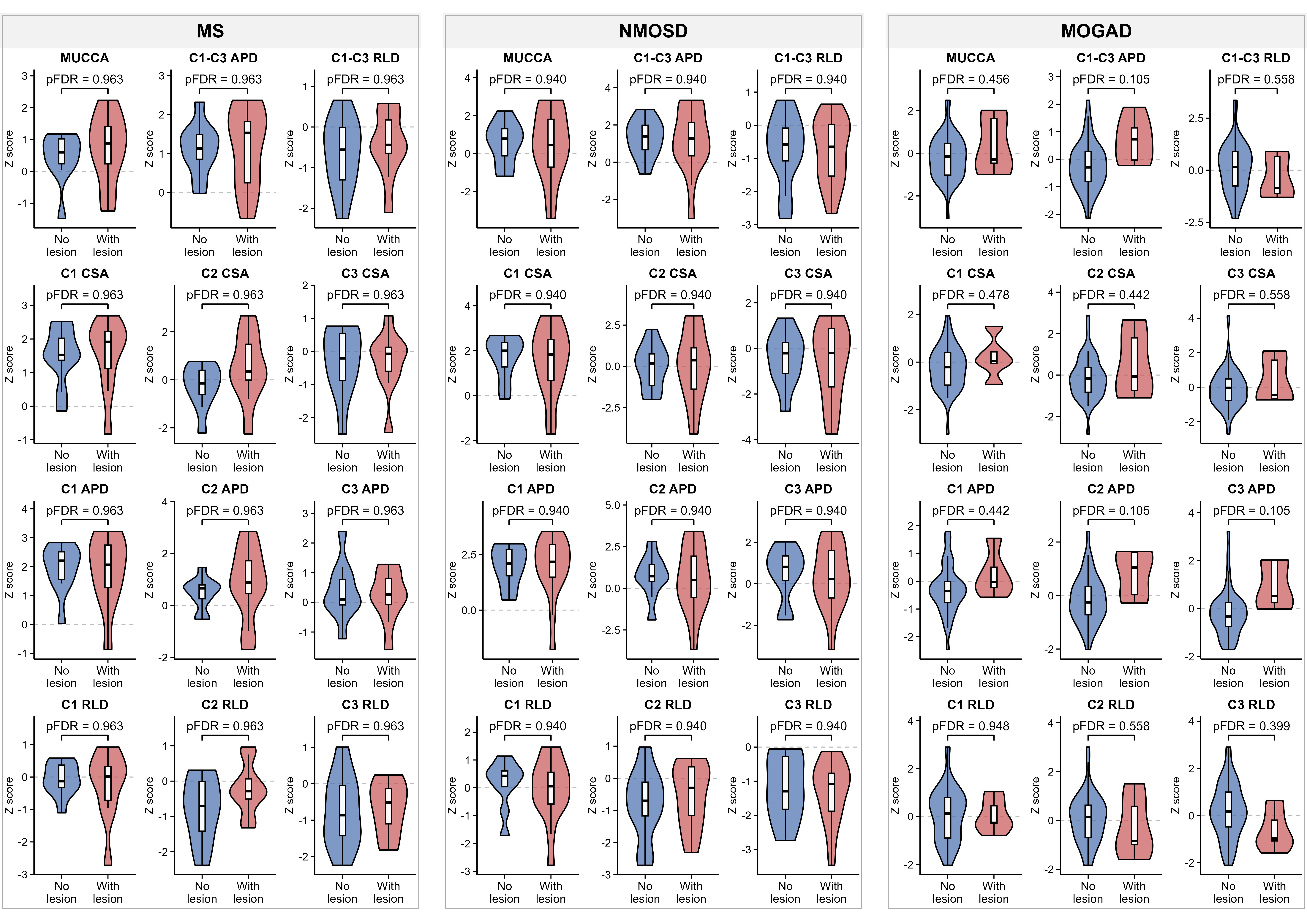
**Fig. S9. Comparison of upper cervical spinal cord morphology between patients with and without MRI-visible spinal cord lesions in MS, NMOSD, and MOGAD.** Violin plots show the distributions of Z scores for 12 upper cervical spinal cord morphological measures in patients with both T1WI and T2WI available across MS, NMOSD, and MOGAD, stratified by the presence or absence of MRI-visible spinal cord lesions. Embedded box plots indicate the median and interquartile range. Group differences were assessed using an independent-samples t-test or Welch’s t-test when normality assumptions were met, and the Mann–Whitney U test otherwise. P values were adjusted for multiple comparisons using the Benjamini–Hochberg FDR method separately across the 12 measures within each diagnostic group. Adjusted P values are shown as pFDR.

Note: MS, multiple sclerosis; NMOSD, neuromyelitis optica spectrum disorder; MOGAD, myelin oligodendrocyte glycoprotein antibody-associated disease; MUCCA, mean upper cervical cord area; CSA, cross‑sectional area; APD, antero-posterior diameter; RLD, right-left diameter; FDR, false discovery rate.

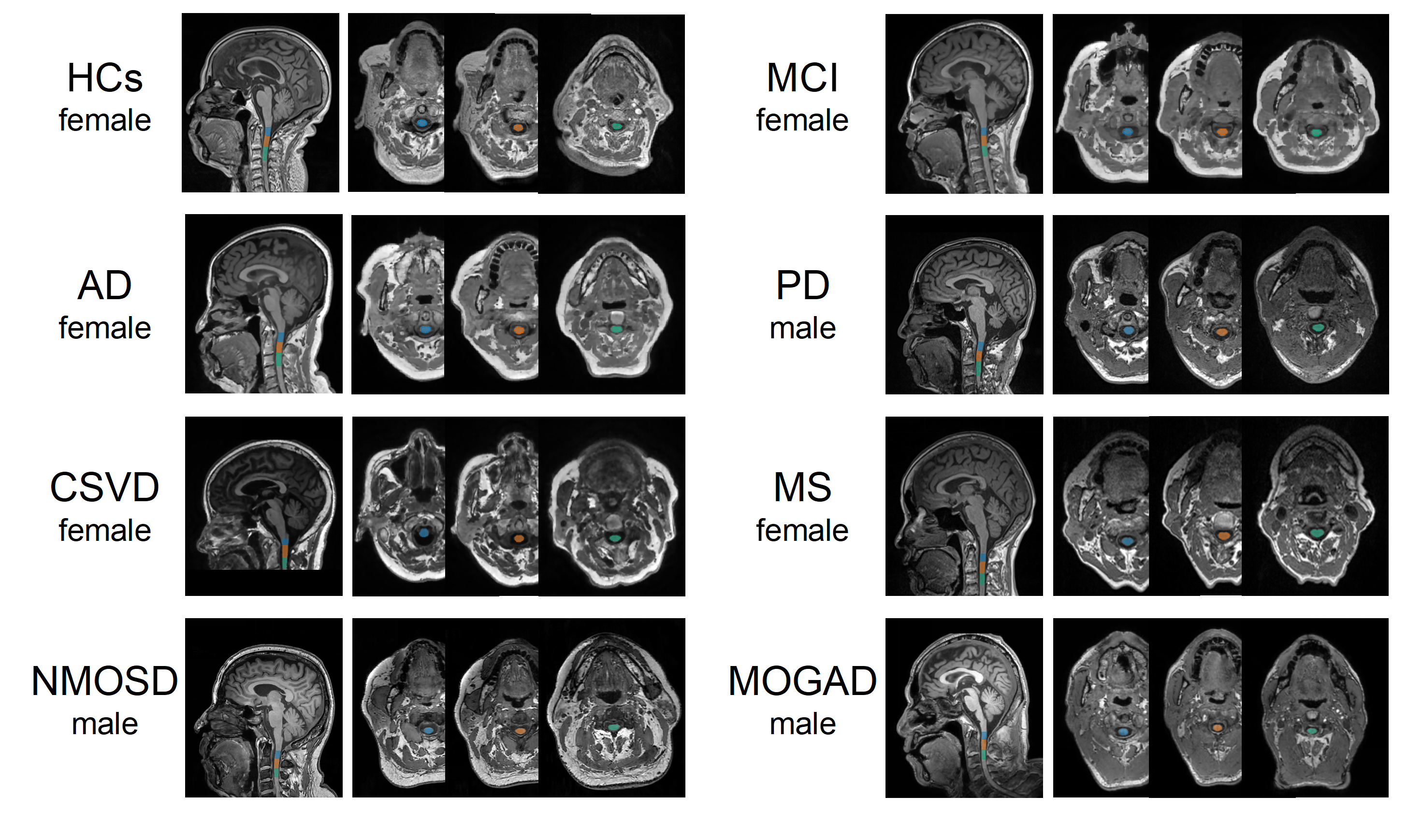

**Fig. S10. Representative upper cervical spinal cord segmentation across diagnostic groups.** Representative images illustrate segmentation of the upper cervical spinal cord from C1 to C3 in individuals from different diagnostic groups. Each panel represents one participant, with the corresponding diagnosis indicated.

Note: HCs, healthy controls; MCI, mild cognitive impairment; AD, Alzheimer’s disease; PD, Parkinson’s disease; CSVD, cerebral small vessel disease; MS, multiple sclerosis; NMOSD, neuromyelitis optica spectrum disorder; MOGAD, myelin oligodendrocyte glycoprotein antibody-associated disease.

**Table S1. Available MRI scanners and acquisition parameters**

| **Scanner** | **Sequence** | **TR/TE (ms)** | **Flip angle (°)** | **Matrix sizes** | **Voxel size (mm)** |
| --- | --- | --- | --- | --- | --- |
| Siemens Skyra | MPRAGE | 2300/2.3 | 8 | 256×256×192 | 1.0×1.0×1.0 |
| Siemens Prisma | MPRAGE | 2400/2.9 | 8 | 256×256×176 | 1.0×1.0×1.0 |
| Siemens Trio | MPRAGE | 1900/2.3 | 9 | 256×256×176 | 1.0×1.0×1.0 |
| GE Premier | SPGR | 8.2/3.2 | 12 | 256×256×176 | 1.0×1.0×1.0 |
| GE Discovery MR 750 | SPGR | 8.2/3.2 | 9 | 256×256×188 | 1.0×1.0×1.0 |
| Philips Ingenia CX | MPRAGE | 7.0/3.0 | 8 | 256×256×196 | 1.0×1.0×1.0 |
| UIH uMR 880 | FSPGR | 7.8/3.2 | 12 | 256×256×196 | 1.0×1.0×1.0 |

Note: MPRAGE, Magnetization Prepared Rapid Gradient Echo; SPGR, Spoiled Gradient Recalled Echo; FSPGR, Fast Spoiled Gradient-Recalled Echo.
